# A New State-Space Epidemiological Model for Cost-Effectiveness Analysis of Non-Medical Interventions- A Study on COVID-19 in California and Florida

**DOI:** 10.1101/2020.12.24.20248830

**Authors:** Vishal Deo, Gurprit Grover

## Abstract

In the absence of sufficient testing capacity for COVID-19, a substantial number of infecteds are expected to remain undetected, and hence, are not quarantined. This, in turn, defeats the whole purpose of non-medical containment measures, like quarantine, lockdown, travel ban, physical distancing *etc*., by keeping the average reproduction rate above 1. To stress upon the importance of extensive random testing for breaking the chains of transmissions, we have formulated a detailed framework for carrying out cost-effectiveness analysis (CEA) of extensive random testing in comparison to targeted testing (the existing testing policy followed by most countries). This framework can be easily extended for CEA of any other non-medical or even medical interventions for containing epidemics.

We have developed a new version of the basic susceptible-infected-removed (SIR) compartmental model, called the susceptible-infected (quarantined/ free) - recovered-deceased [SI(Q/F)RD] model, to incorporate the impact of undetected cases on the transmission dynamics of the epidemic. Further, we have presented a Dirichlet-Beta state-space formulation of the SI(Q/F)RD model for the estimation of its parameters using posterior realizations from Gibbs sampling procedure. As an application, the proposed methodology is implemented to forecast the COVID-19 transmission in California and Florida, and further carry out CEA of extensive random testing over targeted testing.

**Highlights:**

- Estimated values of excess deaths associated with COVID-19 are used to account for underreporting, and for calibrating data to obtain actual counts of cases.
- A new flexible version of SIR compartmental model, called SI(Q/F)RD, is introduced to facilitate in the CEA exercise.
- Dirichlet-Beta state-space formulation of the SI(Q/F)RD model is used to predict the transmission dynamics of the epidemic.
- CEA is conducted in terms of outcome (reduction in infections and deaths) and total cost of tests.
- Proposed methodology is applied on the data of California and Florida.

## 1. Introduction

Whenever we encounter an epidemic, the best medical intervention we can think about for containing its spread is a quick resort to mass vaccination of the susceptible population. However, when we face a pandemic like COVID-19, caused by the SARS-CoV-2 virus, the novelty of the virus puts up an arduous challenge before the scientists to develop an effective vaccine in a short span of time. Further, the necessary safety protocols underlining the testing and approval of vaccines, followed by the herculean task of manufacturing it in abundance, makes it practically impossible to get a potent vaccine within a year of the outbreak of the pandemic. Consequently, it is of paramount importance to strategically implement non-medical interventions, like physical distancing, quarantine, lockdown measures *etc*., to minimize the spread of the infection. The rationale behind these non-medical containment measures is to break the chain of infections by bringing down the basic reproduction number/ rate, *R*_*0*_, below one. *R*_*0*_ is an important factor for risk assessment of any epidemic and is defined as the expected number of secondary cases that arise from a typical infectious index-case in a completely susceptible host population. When *R*_*0*_ is less than one, an infected case is expected to produce less than one new infected. This marks the decline in the number of infecteds over time and, eventually, the epidemic dies out.

As per the scientific brief of the World Health Organization (WHO) published on its website on 9 July 2020, transmission of SARS-CoV-2 occurs primarily between people through direct, indirect, or close contact with infected people through infected secretions such as saliva and respiratory secretions, or through their respiratory droplets, which are expelled when an infected person coughs, sneezes, talks or sings; refer WHO (2020 a). That is, in order to break the chains of transmissions of SARS-CoV-2, the objective of the preventive measures should be to minimize the contact of susceptibles with infected people. The first step towards this goal is to identify the infecteds so that they can be kept in quarantine till they are no longer infectious. However, high variability in the level and the nature of symptoms in infecteds, coupled with a significant length of incubation period, poses a difficult challenge to frame a targeted testing strategy which can serve the purpose effectively. Although contract tracing can help in identifying the chains of transmission associated with detected cases, presence of a high proportion of asymptomatic cases [Byambasuren *et al*. (2020), CDC (2020 a)], also capable of transmitting infection, flags concerns about the reliability of the strategy. Further, in the presence of high proportion of asymptomatic cases, limiting testing to the symptomatic individuals will also fail to serve the objective of detecting and quarantining all infecteds. So, apart from testing symptomatic individuals (mild or severe), and identifying and testing high risk individuals having history of contact with infected people, the situation demands aggressive random testing to isolate even the asymptomatic cases from the population. In the absence of adequate amount of random testing, a significant number of infecteds, especially asymptomatic individuals, may remain undetected (*i*.*e*., number of cases will remain significantly underreported). Since the undetected cases are not quarantined, they are expected to remain infectious in the population for a relatively much longer period as compared to those who are detected and quarantined. Lack of any visible symptom in the undetected cases increase the likelihood of susceptibles spending prolonged period in their close proximity. Thus, the undetected cases can exhibit a strikingly higher reproduction rate as compared to that of their quarantined counterparts. To be precise, aggressive random testing is imperative towards fulfilling the objective of breaking the chains of transmissions of SARS-CoV-2 using non-medical containment measures.

Despite repeated appeals and advisories to all countries from the WHO to employ extensive random testing, only few countries have shown intentions to conduct adequate number of COVID-19 tests [WHO (2020 b)]. Rate of positive tests at a place can give an indication about the adequacy, or inadequacy, of the number of tests being carried out in the region. New Zealand has set an extraordinary example in quickly containing the epidemic by efficiently adhering to the strategy of aggressive testing and isolation of infecteds to break the chains of infection. As on 25 August 2020, New Zealand has reported a total of 1690 confirmed cases of which 1539 have recovered, 129 are active and 22 have died. These are very encouraging figures, especially when compared with those of the countries like USA, Brazil, UK, India, Russia and many others. Till 19 August 2020, the overall percentage of positive tests in New Zealand is reported to be 0.23% [Ministry of Health (2020)]. While in USA, the percentage of positive tests has remained quite high since the beginning, with the overall positive percentage staring at 9%. Also, there is a lot of variation between the percentages of positive tests reported by different states in USA, which varies between 0.53% (Vermont) to 100% (Washington) as reported on 26 August 2020 by the Johns Hopkins University [JHU (2020)]. As per the recommendation of the WHO, the rates of positivity in testing should remain at 5% or lower for at least 14 days at a stretch before governments decide about relaxing the containment/ lockdown measures. This advisory from the WHO is based on the rationale that very high positive rates may indicate that people with only severe symptoms are getting tested and all the asymptomatic cases or cases with mild symptoms are being left out. That is, a high rate of positivity may imply that the testing capacity of the state/ country is insufficient to gauge the actual size of the outbreak, leading to a significant amount of underreporting of cases.

The above discussion stresses on the fact that the success of any non-medical containment measure relies heavily on the ability to have sufficient testing capacity to identify and isolate the infected people. Even the strongest of the lockdown measures will fail to serve its purpose of breaking transmission chains unless it is accompanied with sufficient amount of random testing. Or, in other words, in the absence of sufficient testing capacity, lockdown measures can only succeed in delaying the spread of the epidemic. Citing these reasons, we have considered analysing the effectiveness of extensive random testing over targeted testing as a non-medical intervention in containing the spread of COVID-19-both in terms of effectiveness in reducing transmission rates and the associated costs. By the phrase ‘targeted testing’ we imply testing of only symptomatic and high-risk people. To perform the cost-effectiveness analysis (CEA), we have considered the case of two of the worst affected states of USA, California and Florida, which have very high percentages of positive tests. Since the level of testing, and protocols/ procedure of reporting of number of deaths vary between different state jurisdictions, the level of underreporting of deaths and cases can also be expected to vary between states. This is the reason that we have performed state-wise analyses, rather than analysing the combined data of the USA. For forecasting the transmission dynamics of the pandemic under different assumptions regarding prevalence of underreporting, we have developed a new extension of the state-space SIR model given by Osthus *et al*. (2017). It should be noted that, although underreporting of cases can occur because of various other reasons, like poor communication between state and hospitals, conscious data manipulation to conceal failures of administration, anomalies in protocols used for declaring epidemic related deaths, lack of proper digital infrastructure to keep reliable records, to name a few, we have assumed that lack of sufficient testing is the sole (or at least major) reason for underreporting. This assumption definitely holds for a developed country like USA, where other reasons like lack of proper communication or digital infrastructure can be conveniently crossed off.

## 2. Methodology

To realize the objective of conducting CEA of extensive random testing against targeted testing, we propose the following sequence of steps, which are then implemented on the COVID-19 time-series data of California and Florida.

### 2.1. Estimation of underreporting of deaths due to COVID-19

Underreporting of deaths has been estimated using the concept of excess deaths. Excess deaths due to an epidemic (COVID-19 in our case) can be estimated as the difference between the total number of deaths reported in the period of epidemic (from all causes) and the expected number of baseline deaths due to all other causes in the absence of COVID-19. One popular method to calculate the expected number of baseline deaths in the absence of COVID-19 is to fit a Poisson regression to the time-series (weekly) data of death counts, and then projecting the baseline death counts till the required future point in time. An over-dispersed Poisson generalized linear models with spline terms is used to model trends in counts, accounting for seasonality [CDC (2020 b), Weinberger *et al*. (2020), Rivera *et al*. (2020)]. The model is also adjusted for year-to-year baseline variation and any pre-existing epidemic, like influenza epidemic. For our study we have used weekly estimates of excess deaths published by the Centers for Disease Control and Prevention (CDC) for the two states under consideration for the CEA [CDC (2020 b)]. The estimated excess death counts in the presence of COVID-19 are taken as the estimates of actual number of deaths due to the pandemic. The reported daily deaths due to COVID-19 are summed over weeks to find weekly reported deaths. The difference between the weekly excess deaths and weekly reported deaths give us the estimate of the unreported deaths due to the pandemic. For further analysis, the weekly estimate of unreported deaths due to COVID-19 is distributed among each day of the corresponding week as per the proportion of the number of pandemic related deaths reported on a day out of the total number of deaths due to the pandemic reported in that week. If all days of a week have zero reported deaths, the total number of unreported deaths is equally distributed among all seven days. Combining the additional death counts assigned to each day to the already reported death counts for the day gives us the calibrated daily time-series data on actual number of deaths due to COVID-19. The calibrated data is smoothed using the method of LOESS regression before proceeding with further calculations.

### 2.2. Estimation of underreporting of number of infecteds and total confirmed cases

Actual daily number of infecteds can be estimated using a reliable estimate of case fatality rate (CFR). As the reported data on number of cases is expected to suffer from underreporting, the CFR estimated on the basis of the population level data will be misleading. If we assume that the level (or proportion) of underreporting of deaths and infecteds are same, the CFR estimated from the reported data will be a reliable estimate of the true CFR. However, this is hardly the case observed in reality, and the proportions of underreporting of deaths and infecteds usually differ considerably. An estimate of CFR based on a (follow-up) individual patient level data is deemed as most reliable [Atkins *et al*. (2015)]. So, a simple rule of thumb to know if the levels of underreporting of deaths and infecteds can be assumed to be same is to compare the delayed CFR obtained from reported data with the one obtained from the individual patient level data. If they vary significantly, we should assume that the levels of underreporting are different for number of deaths and number of infecteds. In such a situation, it is advisable to use the CFR obtained from the individual patient level data as the best estimate for the true CFR of the epidemic. Once a reliable estimate of the CFR is obtained, it can be used to calibrate the data for the number of true infecteds on each day using the estimated counts of true deaths and the average delay between infection and death. That is, if the average duration between infection (or detection/ reporting of infection) and death associated with COVID-19 is known to be, say, *h* days, and suppose that *D*_*t*_ number of people have died of the infection on a particular day *t*, then *I*^*C*^_*t-h+1*_ *=* (*D*_*t*_ */* CFR) number of new cases are expected to be actively infected *h* days prior to the day of death.

If daily reported data on the number of recovered cases (***R***_***t***_) is available, it can be inflated according to the proportion of underreporting estimated in the number of infecteds as follows.

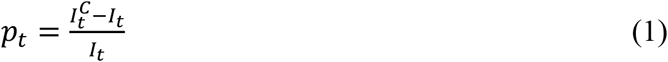

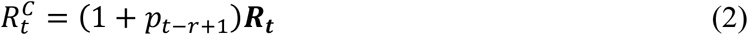

where, *p*_*t*_ is the estimated proportion of underreporting of infecteds at time *t*, 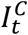 is the calibrated value of number of infecteds at time *t, I*_*t*_ is the reported value of number of infecteds at time *t*, 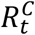 is the calibrated value of number of recovered cases at time *t*, ***R***_***t***_ is the reported number of recovered cases at time *t* and *r* is the average duration from infection to the recovery of patients. If the daily number of recovered cases is not reported, or if it is not reliable, a fairly good calibration can be done using *r* and (1-CFR). Calculations will be similar to that used for the calibration of number of infecteds. Further, sum of daily calibrated data of total infecteds, total recovered and total deaths will give us the estimated values of the total number of confirmed cases.

### 2.3. The proposed SI(Q/F)RD epidemic model

This extension of the popular Susceptible-Infected-Removed (SIR) compartmental model is designed to account for underreporting and its impact on the trajectory of the epidemic. Here, SI(Q/F)RD stands for Susceptible-Infected (Quarantined/ Free)- Recovered-Deceased model. Underreporting is assumed to be the result of some infected cases not getting detected because of the lack of adequate testing capacity. That is, a proportion of the infecteds are detected (*p*) and quarantined (mostly symptomatic cases), while the rest of the infecteds (mostly asymptomatic cases) are undetected and roam freely among the susceptibles. This leads to the belief that the undetected cases are expected to infect the susceptibles at a higher rate (*β*_*2*_), than that of their quarantined counterparts (*β*_*1*_). The proportion of detected cases, *p*, can vary with time if testing capacity is increased or decreased over the period of the epidemic, and can be taken as a function of time *t*, say, *p*_*t*_. The overall structure of this model is presented in Figure 1. Since the quarantined infecteds consist of symptomatic cases, including critical cases, quarantined infecteds can be expected to be at a higher risk of death on an average. Consequently, different death rates are assumed for quarantined and undetected cases. Although there is no strong logic behind choosing different average recovery rates for quarantined and undetected cases, different recovery rates can be assumed if any scientific evidence exists in its favour. Otherwise, we can assume that both groups of infecteds have equal average recovery rate.

**Figure 1:**
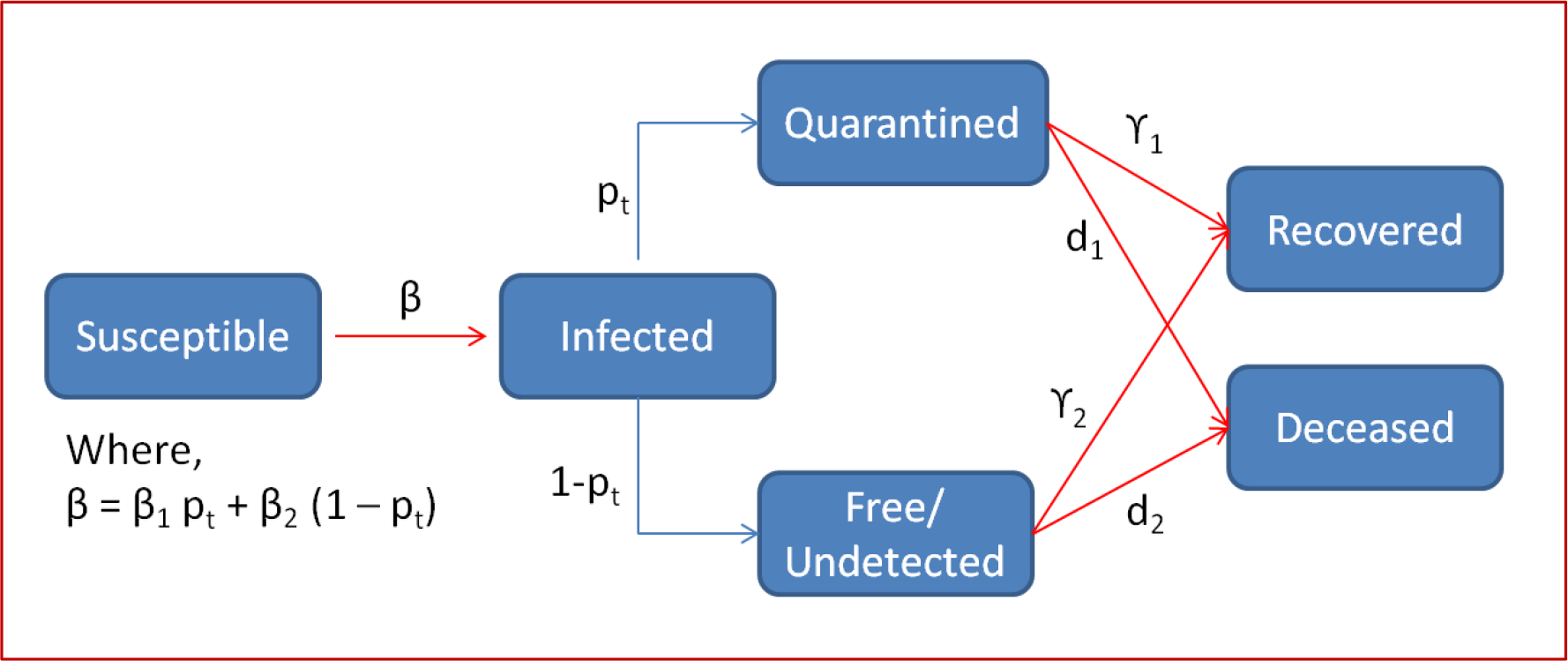
SI(Q/F)RD model structure- *p*_*t*_ is the proportion of infecteds detected and quarantined, *1-p*_*t*_ is the proportion of infecteds who are undetected and roaming freely among the susceptibles, *β*_*1*_ is the transmission rate associated with quarantined infecteds and *β*_*2*_ is the transmission rate associated with undetected infecteds, *γ*_*1*_ and *d*_*1*_ are rate of recovery and rate of death for quarantined cases and *γ*_*2*_ and *d*_*2*_ are rate of recovery and rate of death for undetected cases.

The set of differential equations quantifying the transitions defined in Figure 1 can be expressed as follows.

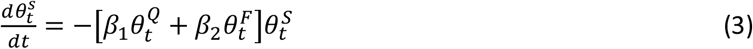

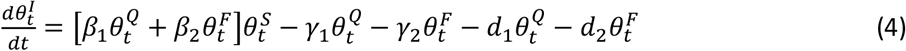

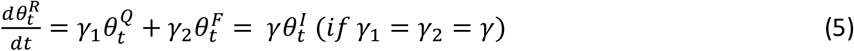

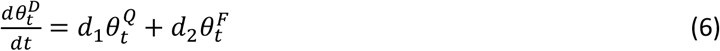

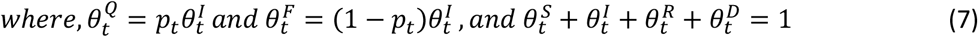

Here, 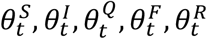 and 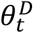 are the true but unobserved (latent) prevalence of susceptibles, infecteds, infected and quarantined, infected and free (undetected), recovered, and deceased respectively. In other words, they are the probabilities of a person being in the respective compartments at time *t*. Also, let 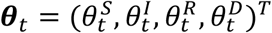 be the latent population prevalence. Solution of this set of differential equations can be obtained using Runge-Kutta approximation. Let *f* (***θ***_*t*−1_, ***β, γ, d***) denotes the solution of the set of differential equations for time *t*, where the function takes the values of the vectors ***θ***_*t*−1_, ***β*** = (*β*_1_, *β*_2_)^*T*^, ***d*** = (*d*_1_, *d*_2_)^*T*^and ***γ*** = (*γ*_1_, *γ*_2_)^*T*^ as the arguments. Then the fourth order Runge-Kutta approximation for the solution of these differential equations can be expressed as follows.

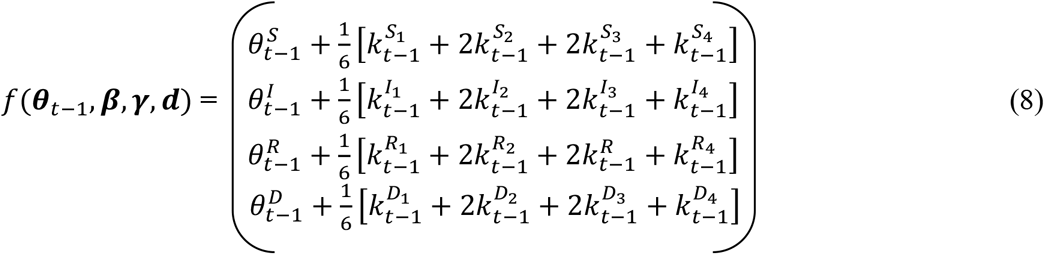

where,

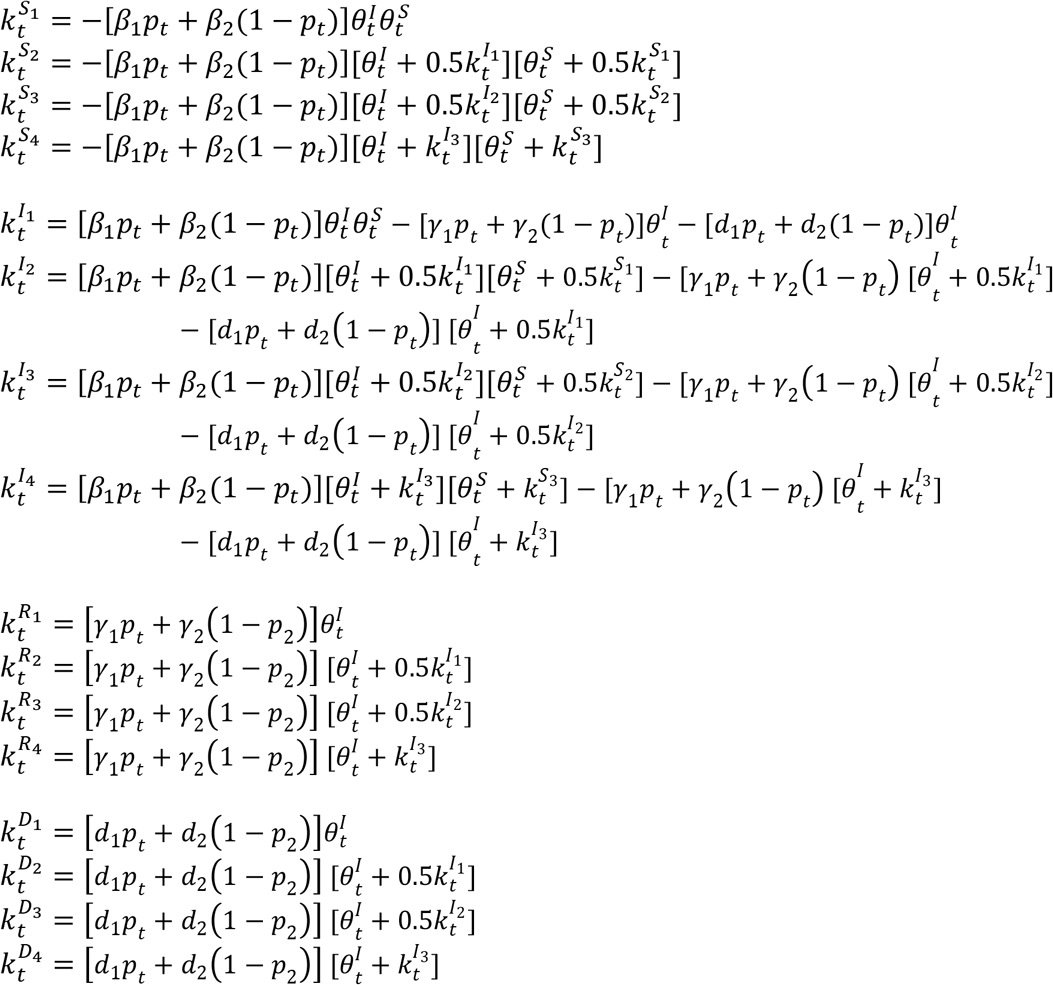

### 2.4. Dirichlet-Beta state-space formulation of the SI(Q/F)RD model

To account for the uncertainties in the epidemiological parameters and the transmission dynamics of the epidemic, we define a flexible state-space probabilistic model based on the deterministic SI(Q/F)RD model. Osthus *et al*. (2017) introduced a Dirichlet-Beta state-space model based on the basic SIR model. We have extended the Dirichlet-Beta state-space model in accordance with the SI(Q/F)RD structure to estimate the model parameters, and forecast the transmission dynamics of the epidemic. Let 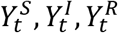 and 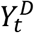 be the observed proportion of susceptibles, infecteds, recovered and deceased respectively. Then the Bayesian hierarchical state-space SI(Q/F)RD model can be defined as follows.

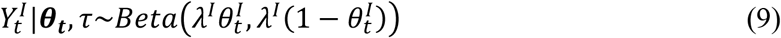

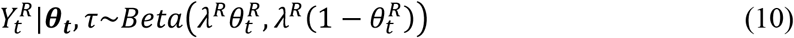

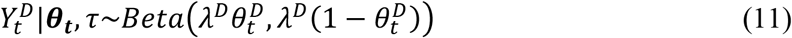

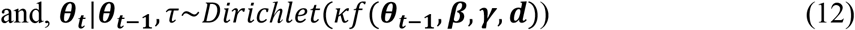

where, *τ*= {***θ***_**0**_, *κ*, ***β, γ, d***,*λ*^*I*^,*λ*^*R*^,*λ*^*D*^}, ***θ***_**0**_ is the baseline value of the vector ***θ***_***t***_, and *λ*^*I*^,*λ*^*R*^,*λ*^*D*^, **κ** > 0 control the variances of the distributions defined in equations (9), (10), (11) and (12) respectively. All other notations are as they are defined in section 2.3. From equation (12) it is apparent that ***θ***_***t***_, *t* = 1,2 …, *T*, is a first-order markov chain. Also, the equations (9), (10) and (11) suggest that, for *t* ≠ *s*, 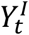 is independent of 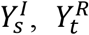 is independent of 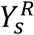, and 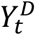 is independent of 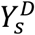, given 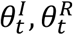 and 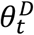 respectively.

Prior distributions of the model parameters can be defined as follows.

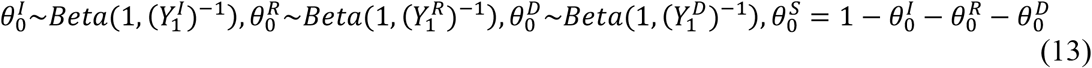

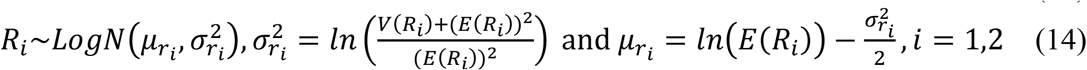

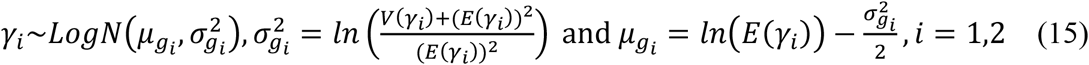

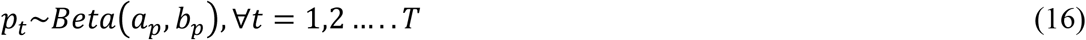

*R*_*1*_ and *R*_*2*_ are basic (average) reproduction rates associated with quarantined (Q) and undetected (F) infecteds, respectively. That is, 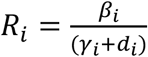, *i* = 1,2.

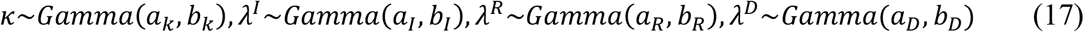

The hyperparameters of these Gamma prior distributions can be assumed according to the size of variability to be allowed in the Beta and Dirichlet distributions defined in equations (9)-(12). The higher the values of the parameters *κ,λ*^*I*^,*λ*^*R*^ and*λ*^*D*^, the lower will be the variance of the respective Beta and Dirichlet distributions. If limited prior information is available regarding these parameters, a relatively flat Gamma prior distribution with a high expected value and a relatively higher variability is assumed while choosing the values of the hyperparameters *a’s* and *b’s*. Hyperparameters of the prior distribution of *p*_*t*_ are obtained from method of moments using the mean and variance of the daily estimates of underreporting. Hyperparameters of the lognormal distributions defined in the expressions (14) and (15) can be either based on historical knowledge on a similar epidemic, or can be estimated on the basis of the observed data. In our study, we have presented a time-series SIR (TSIR) model based technique to estimate the hyperparameter for transmission rate, ***β***. Values for the hyper parameters ***γ*** and ***d*** are calculated using required information from published literature on COVID-19. *γ*_1_ = *γ*_2_ = *γ* (*say*) is taken as the inverse of the average recovery period. We have assumed equal recovery rates for both groups, quarantined and undetected, as there is no scientific evidence to suggest that there can be significant difference in the average recovery time of the two groups. The components of ***d***, *viz*., *d*_*1*_ and *d*_*2*_ are estimated as, 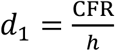 and 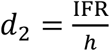, where *h* is the average number of days from infection till death and IFR is the infection fatality rate. While CFR is the ratio of the number of deaths divided by the number of confirmed cases of disease, IFR is the ratio of deaths divided by the number of actual infections with SARS-CoV-2 and is generally expected to be lower than CFR.

### 2.5. Estimation of the hyperparameter *β* using TSIR model

The two components of the vector ***β*** = (*β*_1_, *β*_2_)^*T*^ have to be estimated separately, such that they conform to their definitions. To do so, we have made some assumptions about the reported data, based on certain practical considerations. If it is known that there was no significant effort to quarantine infecteds or to promote physical distancing during the initial period of the epidemic, the transmission rate observed during that period can be safely considered as an initial estimate of *β*_2_ (the transmission rate due to infecteds who are not quarantined). Once the containment measures are imposed, the transmission rate based on the reported data is expected to change (reduce) and average transmission rate observed over the entire period of reporting can be considered as a safe initial estimate (hyperparameter estimate) for *β* (the overall average transmission rate as a result of both quarantined and undetected infecteds). This logic can be implemented using the TSIR model to estimate the hyperparameters as follows.

In TSIR model, the response, being a count variable, is assumed to follow certain discrete count process distribution like Poisson distribution or Negative Binomial distribution. The basic structure of TSIR model can be defined as follows [(Bjørnstad *et al*. (2002), Finkenstadt *et al*. (2002), and Grenfell *et al*. (2002)].

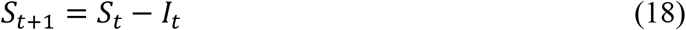

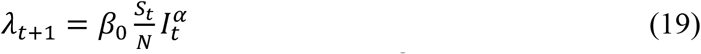

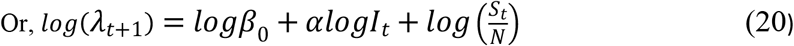

where, *S*_*t*_ and *I*_*t*_ are the number of susceptibles and infecteds (or infectives) at time *t, N* is the population size, *β*_*0*_ is the transmission rate and *λ*_*t*+1_ is the expected number of new infecteds at time *t+*1. New number of infecteds is assumed to follow Negative Binomial (or Poisson) distribution and a generalized Negative Binomial (or Poisson) linear model with log link is fitted with *logI*_*t*_ as a covariate and 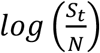 as an offset variable. The exponent *α* is expected to be just under 1 (*i*.*e*., close to 1) and is meant to account for discretizing the underlying continuous process. However, we use an alternative interpretation of *α* based on the basic SIR model defined in equation (21). This method is drawn from our prior work where we have proposed a new method for obtaining time-varying estimates of transmission rate using TSIR model [Deo *et al*. (2020)]. It is to be noted that the transmission rate is assumed to be time-varying, and hence, denoted as *β*_*t*_.

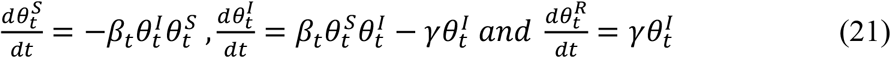

Using (21), the expression for expected number of new infecteds at time *t+*1 (taking *α* = 1) with a time-varying transmission rate can be written as follows.

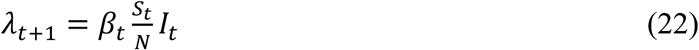

Comparing equations (19) and (22), we can see that if *α* = 1 (or close to 1), *β*_*t*_ *= β*_*0*_ (constant over time). However, if the value of *α* deviates considerably from 1, it has impact on the effective value of transmission rate, thus making the effective rate of transmission time-dependent. That is, in such cases *α* assimilates the empirical changes in transmission rate over time. Further, using equations (19) and (22), we can write,

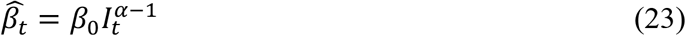

Now, suppose *T*_*1*_ represents the initial period of the epidemic, when proper quarantine protocols were not in place, and *T* represents the entire period for which the reported data on the epidemic is available. The estimates of *α* and *β*_*0*_ obtained from the fitting of the TSIR model shall be used in equation (23) to find estimates of transmission rate at each time *t*, say 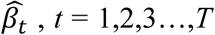. Average of these estimates over a time period will give us the estimate of average transmission rate for that period. That is, the estimates of the transmission rates will be taken as, 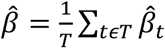 and 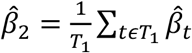. Then a reliable estimate of *β*_1_, the transmission rate due to the infecteds who are quarantined, can be obtained using the relation, 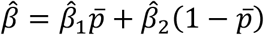; *where* 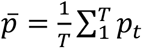. As a simpler but logical alternative to this step for finding 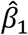, we can make use of the fact that the infecteds who are quarantined are expected to spread infection for approximately only one-third of the duration as compared to those who are not quarantined. This estimation is based on the fact that quarantined patients spread infections mostly in the incubation period of around 4-5 days, prior to getting quarantined, while infecteds who are not quarantined are expected to freely spread infection for the entire average infectious period of 14 days. So, after estimating 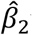 using the method described above, we can take 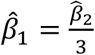, as the initial estimate.

This exercise forms an essential part of the overall methodology presented in our study for conducting the CEA. This is because, the utility of extensive testing and subsequent quarantining of infecteds can be assessed only when we can estimate the changes in transmission rate expected to be brought about by these measures.

### 2.6. Estimating parameters of the state-space SI(Q/F)RD model and forecasting

Posterior realizations on the parameters of the state-space model are generated using Gibbs sampling MCMC approach. The model is adopted in JAGS format and implemented in R programming using the package R2jags. Mean of posterior realizations of a parameter is taken as the posterior estimate of the parameter. Further, 0.025 and 0.975 quantiles of the posterior realizations are taken as the 95% credible intervals (CI) of the posterior estimates. Let *t*_*0*_ be the time till which the observations are available, and suppose that we wish to forecast the values of observed process 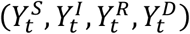 from *t*_*0*_+1 till the time *T*. We follow the following iterative steps to achieve our goal.

a. We generate *L* posterior realizations on the latent prevalence process 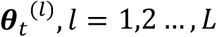 using Gibbs sampling approach, at each time point *t* = *t*_*0*_ +1, *t*_*0*_ + 2…..,*T*. Here *L* is a sufficiently large number, say 1000 or more.
b. At each *t* (= *t*_*0*_ +1, *t*_*0*_ + 2…..,*T*), and at each posterior realization of the prevalence process ***θ*** ^(*l*)^, *l* = 1,2 …, *L*, values of the observed process, say 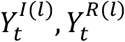 and 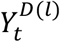 are simulated from their conditional distributions, 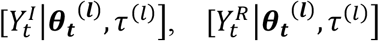 and 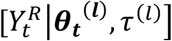, which are defined in the equations (9), (10) and (11), respectively. Further, using the posterior realizations of 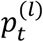, at each *l* and each *t*, 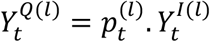 and 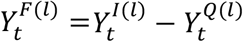 are also obtained. At each *t*, mean of the *L* simulated values serves as the estimate (forecasted value) of the respective variable (compartment proportion). 95% credible interval of each variable, at each time t, is also obtained using the 0.025 and 0.975 quantiles of the *L* values.

### 2.7. Cost-effectiveness analysis

The two states of USA chosen for our study, California and Florida, have very high rates of positivity of tests. It indicates that instead of resorting to sufficient amount of random testing, these states may have relied on targeted testing, *i*.*e*., testing of only symptomatic and high-risk individuals. Very high CFR rates in these states also corroborate the possibility of insufficient testing, and hence, underreporting of cases. Thus, as mentioned earlier in the introduction section, our objective is to conduct CEA of extensive random testing aimed at isolating almost all infecteds, as recommended by the WHO, over targeted testing which is seem to be the strategy followed by the states. Extensive random testing can be expected to result in a significant rise in expenditure on the testing kits and medical personnel. However, it can play a major role in breaking the chains of transmission and hence, result in a significant reduction in the overall number of infecteds and deaths due to the COVID-19 epidemic. The outcome of CEA will tell us how much additional overall cost is required to save one additional person from getting infected, or to save one additional person from dying due to the infection. That is, CEA will be conducted in terms of the outcomes, ‘infection’ and ‘death’. It should also be noted that, if the total duration of the epidemic is reduced drastically because of the recommended intervention ‘extensive random testing’, the overall expected cost may even come out to be lesser than the expected cost of using targeted testing strategy.

The methodological steps mentioned in the sections 2.4, 2.5, and 2.6 are associated with estimation and forecasting under the actual scenario based on the observed data. That is, the results obtained from these steps will pertain to the base intervention of ‘targeted testing’ which is being currently followed by the two states. To derive the outcomes pertaining to the recommended intervention, i.e., ‘extensive random testing’, following procedure is followed.

a. In terms of the SI(Q/F)RD model, the major difference between the outcomes of the two scenarios will rely on the difference in the proportion of infecteds being detected and quarantined, *i*.*e*., *p*_*t*_.
b. It will be impractical to assume that 100% infecteds can be detected using extensive random testing. This is because even popular tests like the reverse transcription polymerase chain reaction (RT-PCR), which is also recommended by the WHO [WHO (2020 c)], do not have 100% sensitivity and specificity. Sensitivity and specificity may vary according to the laboratory settings and expertise levels of the medical practitioners. Different studies have reported varying levels of sensitivity and specificity of the RT-PCR test, mostly ranging from around 80% to 95% [(West *et al*. (2020), Padhye (2020), Tahamtan and Ardebili (2020)]. On a conservative note, we have assumed that the average proportion of detection of infecteds will be 80%, *i*.*e*., *p*_*t*_ will have a mean of 0.8. Instead of assigning a fixed value to *p*_*t*_, we have assumed *p*_*t*_ to follow Beta distribution to introduce realistic variability in the calculations. The mean of the distribution is taken as 0.8 and its variance is obtained from the results of the state-space model estimated by the method described in section 2.6.
c. To simulate a practically realistic situation, we have assumed that the extensive random testing can be applied only after first 30 days of the outbreak of the epidemic. This is because, extensive random testing requires procurement of testing kits and other logistic arrangements on a large scale, which need some time to be organized. To accommodate this assumption into calculations, the mean value of the distribution of *p*_*t*_ for the first 30 days can be based on the average of posterior estimates of *p*_*t*_ for the first 30 days obtained from the state-space SI(Q/F)RD model. That is, we are assuming that there will not be much difference in the outcomes and costs associated with the two interventions in the initial days of the epidemic.
d. Simulation exercise:
  i. At each *t, t = 1,2,…, T, L* number of values are generated on the parameters *R*_1_, *R*_2_, *γ*_1_ and *γ*_2_ from their distributions defined in the equations (14) and (15). The parameters of these distributions are calculated on the basis of their posterior estimates obtained from the state-space SI(Q/F)RD model. The corresponding values on *p*_*t*_ are simulated from its distribution defined in the previous step c. Fixed values of the death rates, *d*_*1*_ and *d*_*2*_, remain same as those for the state-space SI(Q/F)RD model.
  ii. For each combination *(t, l), t = 1,2,…, T* and *l = 1,2,…,L*, the respective simulated values of the parameters are used in the fourth degree Runge-Kutta approximation of the solution of the set of differential equations of the SI(Q/F)RD model to obtain *f*(***θ***_*t*−1_, ***β, γ, d***), *i*.*e*., the values of the latent process at time *t* as a function of their values at time *t-1*. At the start of the iteration, the initial values of these latent process variables are assigned as the vector ***θ***_0_. The mean of the *L* values of the latent process at a time *t* is taken as its estimate, *i*.*e*., 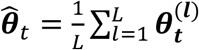. Sample quantiles (0.025, 0.975) are used to obtain 95% credible intervals at each *t*.
  iii. At each *t, t = 1,2,…,T, L* values of *λ*^*I*^,*λ*^*R*^ and*λ*^*D*^ are simulated from their respective Gamma distributions whose parameters are calculated from the posterior estimates of their means and variances obtained from the state-space SI(Q/F)RD model. At each combination (*t, l*), *t = 1,2,…, T* and *l = 1,2,…,L*, using the estimate of the latent prevalence process, 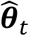, from the previous step and the generated values of *λ*^*I*^,*λ*^*R*^ and*λ*^*D*^, 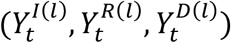 are simulated from their respective Beta distributions. Finally, mean of these *L* values at a time *t* is taken as the estimate of the observed process at *t*. These proportions can be multiplied with the total number of susceptibles (total population of the state) and rounded to obtain the estimated counts of each compartment at time *t, t = 1,2,…T*.
e. Total number of infected cases and total number of deaths, till the end of the epidemic, are calculated from the predictions for each case (interventions). These values give us the difference in outcomes (infection/ death) under two interventions. Let, (*C*_*1*_, *D*_*1*_) be the estimates of total number of infecteds and total number of deaths during the entire course of the epidemic for the base intervention, targeted testing, and (*C*_*2*_, *D*_*2*_) be the respective estimates for the recommended intervention, extensive random testing.

To obtain the estimate of total costs associated with the two interventions we will first need to estimate the total number of tests that will be conducted under the two testing strategies (interventions). For the base intervention of targeted testing, the current percentage of positivity of tests in the state can be used to obtain an estimate of the total number of tests to be conducted by the end of the epidemic. If *r*_*1*_ is the current proportion of positive tests in the state, the estimate of total number of tests which will be conducted under the base intervention will be given as, 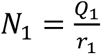, where *Q*_*1*_ is the number of infecteds who are detected and quarantined. For the second intervention of extensive random testing, the proportion of positive tests is taken as the probability that a person in the state got infected during the entire duration of the epidemic and is simply given as, 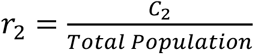. Subsequently, the total number of tests under the second intervention is estimated as, 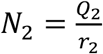, where *Q*_*2*_ is the number of infecteds who are detected and quarantined under the intervention extensive random testing. As an alternative, *N*_*2*_ has also been taken as the total population, assuming that all individuals are tested (once) by the time the epidemic gets over in the state.

Let Z be the per unit average cost of COVID-19 test, then the incremental cost-effectiveness ratio (ICER) is calculated as the ratio of change in cost to the change in outcome as follows,

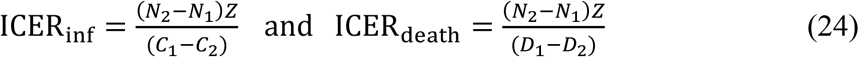

## 3. Implementation and Results

### 3.1. Data

Daily time-series data on total confirmed cases and total deaths for the states of California and Florida are obtained from the github repository of the Centre for Systems Science and Engineering (CSSE), Johns Hopkins University, Maryland, USA [https://github.com/CSSEGISandData/COVID-19]. Daily time-series data till 11 July 2020 was available at the time of procurement of data, and the same has been used for the entire analyses. Data on weekly state-wise estimates of excess deaths associated with COVID-19 till 11 July 2020, calculated as a difference between expected and reported number of deaths from all causes, is obtained from the website of CDC [https://www.cdc.gov/nchs/nvss/vsrr/covid19/excess_deaths.html]. Data on rates of positivity of COVID-19 testing for the two states, California and Florida, are obtained from the official website of Johns Hopkins University on 29 July 2020 [https://coronavirus.jhu.edu/testing/testing-positivity].

### 3.2. CFR and data calibration to account for underreporting

To avoid the initial period of uncertain reporting due to absence of government measures, we have used the data from 02 March 2020 onwards for all analyses. The weekly excess death estimates are used to reconstruct the daily time-series data of deaths using the procedure discussed in section 2.1. According to the results reported by Verity *et al*. (2020) based on patient level data from mainland China, the mean duration from onset of symptoms to death is 17.8 days (95% credible interval 16.9–19.2). They reported the best estimate of case fatality ratio in China as 1.38% (1.23-1.53), with substantially higher CFR in older age groups (0.32% [0.27–0.38] in those aged <60 years *vs* 6.4% [5.7–7.2] in those aged ≥60 years), up to 13.4% (11.2–15.9) in those aged 80 years or older). Their estimate for overall IFR in China is 0.66% [0.39-1.33], with an increasing profile with age. Yang *et al*. (2020) reported that the median time from symptom onset to radiological confirmation of pneumonia is 5 days (interquartile range [IQR] 3–7 days); from symptom onset to intensive care unit (ICU) admission is 11 days (IQR 7–14 days); and from ICU admission to death is 7 days (IQR 3–11 days). That is, the estimate of median time from onset of symptoms to death can be taken as 11 + 7 = 18 days. This estimate of time-to-event of death is consistent with the results of Verity *et al*. (2020). So according to these results, the estimate of average duration from onset of infection to death should be around 17.8 + 5 ≈ 23 days, where 5 days is the average incubation period. However, for estimating delayed CFR based on the reported data and for estimating actual number of infecteds from the calibrated data on number of deaths, we have taken average duration from infected/ reported to death as 18 days. This is because infecteds are generally tested on the onset of symptoms, *i*.*e*., after the incubation period.

The delayed CFR values calculated on the basis of reported data came out to be quite high in both states. For California, the delayed CFR came out to be 3.36%, while for Florida it came out to be 3.17% (Table 1). Both of these estimates are quite higher than the estimate based on individual patient data as reported by Verity *et al*. (2020), *i*.*e*., 1.38%. In fact, some other reports have suggested even lower actual CFR of COVID-19. The Centre for Evidence-Based Medicine (CEBM) at the University of Oxford currently estimates the CFR globally at 0.51%, with all the caveats pertaining thereto; refer [https://www.virology.ws/2020/04/05/infection-fatality-rate-a-critical-missing-piece-for-managing-covid-19/]. Since the estimate of CFR based on follow up data of individual patients is deemed as most reliable, especially during initial stages of the epidemic, as a conservative estimate, we have taken 1.38% as the standard CFR due to COVID-19. Thus, the results of CFR based on the reported data point at underreporting of number of infecteds in both states. Also, significantly high rates of positivity of COVID-19 testing in the two states, 7.47% in California and 18.96% in Florida, as compared to the recommended rate of less than or equal to 5%, indicates lack of adequate amount of random testing. That is, it corroborates our assumption that the current testing strategy employed by the two states largely aim at targeted testing.

**Table 1:** Delayed CFR estimate based on reported data.

| State | Total deaths till<br>16 July 2020 | Total confirmed till<br>29 June 2020 | Average Delayed CFR |
| --- | --- | --- | --- |
| California | 7535 | 223931 | 0.0336 (3.36%) |
| Florida | 4802 | 151389 | 0.0317 (3.17%) |

Using a delay of 18 days (between detection of infection and death), and a CFR of 1.38%, we employ the method discussed in section 2.2 to estimate the actual number of daily infected cases (new cases). It should be noted that due to the lag of 18 days in the formula, number of daily infected cases could be calculated till 24 June 2020 only (18 days prior to 11 July 2020). Data on number of recovered people is not reported for the two states. So, we have calibrated the data for number of recovered cases using the logic that if 1.38% is the average CFR, 98.62% will be the average case recovery rate. Following formula is used to estimate the daily count of recoveries.

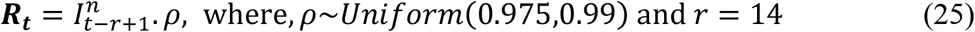

Here,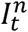 represents new number of infecteds at time *t*. The recovery rate, *ρ*, is assumed to randomly generated between 0.975 and 0.99 to introduce some amount of uncertainty into the calibrated data. The average duration of recovery, *r*, is taken as 14 days based on a WHO report [WHO (2020 d)]. However, at this juncture we should also note that some other studies have reported higher average duration of recovery from COVID-19. Verity *et al*. (2020) have estimated mean duration from onset of symptoms to hospital discharge to be 24.7 days [95% CI: 22.9–28.1]. Recovery time also varies according to the severity of symptoms. Since small recovery time implies faster recovery rate, our choice of 14-day average recovery period (from the onset of symptoms) can be called as a conservative estimate and the forecasts of transmission dynamics based on it can also be expected to be slightly on the conservative side. Since the formula given in equation (25) cannot give us the estimates of the first 13 days, we have constructed the daily recovery data for these initial days using daily recovery rate taken as the inverse of the average duration of recovery. Again, to induce some uncertainty in the data, we have randomly generated recovery rate, *φ*_*t*_, between 0.042 (1/24) and 0.071 (1/14), for each day. Following formula has been applied to implement the idea.

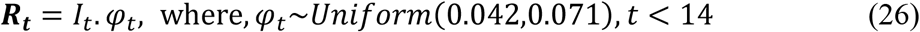

where, *I*_*t*_ is the number of (active) infecteds at time t. For *t < 14*, the already calculated values of recovered cases at time *t + 13* using equation (25), ***R***_***t+13***_, is adjusted by subtracting ***R***_***t***_ from it.

The reconstructed/ calibrated data on number of cases in each compartment is treated as the true data. The ratio of reported number of infecteds and true number of infecteds gives us the estimates of daily proportion of reporting *p*_*t*_ (*Q*_*t*_ *= p*_*t*_. *I*_*t*_). The calibrated data is provided in Appendix-A. Graphs of LOESS smoothed calibrated data on total number of deaths due to COVID-19 for the two states are presented in Figure 2. Summary statistics of *p*_*t*_ are provided in Table 2.

**Table 2:** Summary statistics of *p*_*t*_, proportion of cases reported out of actual number of cases, obtained from the calibrated data.

| Summary | California |  | Florida |  |
| --- | --- | --- | --- | --- |
| $p_t$ values (proportion of reporting) | Mean | Variance | Mean | Variance |
| First 30 days of the observed period | 0.034<br>(3.4%) | 0.0015 | 0.061<br>(6.1%) | 0.008 |
| Last 10 days of the observed period | 0.57<br>(57%) | 0.04 | 0.39<br>(39%) | 0.007 |
| Values lying between the first and the third sample quartiles | 0.26<br>(26%) | 0.01 | 0.23<br>(23%) | 0.007 |

**Figure 2:**
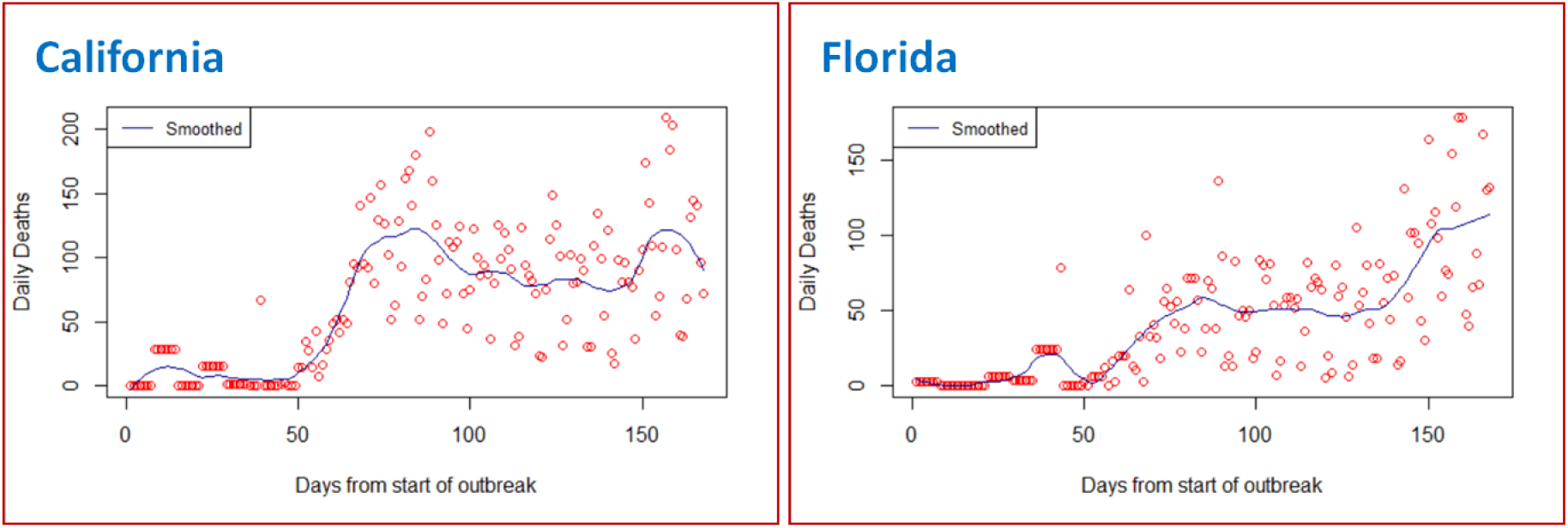
LOESS smoothed calibrated data on total number of deaths due to COVID-19.

### 3.3. Evaluating parameters and hyperparameters of the state-space SI(Q/F)RD model

In California, the first official lockdown measure was implemented on 19 March 2020, while in Florida it was implemented from 01 April 2020. So, the period till 18 March 2020 is considered as initial period of transmission for California, while the period till 31 March 2020 is taken as the initial period of transmission in Florida, for finding initial estimate of the transmission rate in the absence of proper quarantine of infecteds 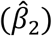. TSIR model described in section 2.5 is fitted assuming both Poisson and Negative Binomial distributions for the count process. Because of lower model deviance, the model with Negative Binomial distribution is chosen over the Poisson model. Models were fitted in IBM-SPSS version 24. The estimated coefficients of the Negative Binomial TSIR models [model defined in equation (20)], for both states, are provided in Table 3. Using these coefficient estimates in equation (23) and taking average over respective initial periods of the COVID-19 epidemic, 02 March 2020-18 March 2020 for California and 02 March 2020-31 March 2020 for Florida, we have obtained the estimates of *β*_2_ for the two states. Initial estimate of *β*_1_ is calculated as one-third of the estimate of *β*_2_. These estimated values are also provided in Table 3.

**Table 3:**
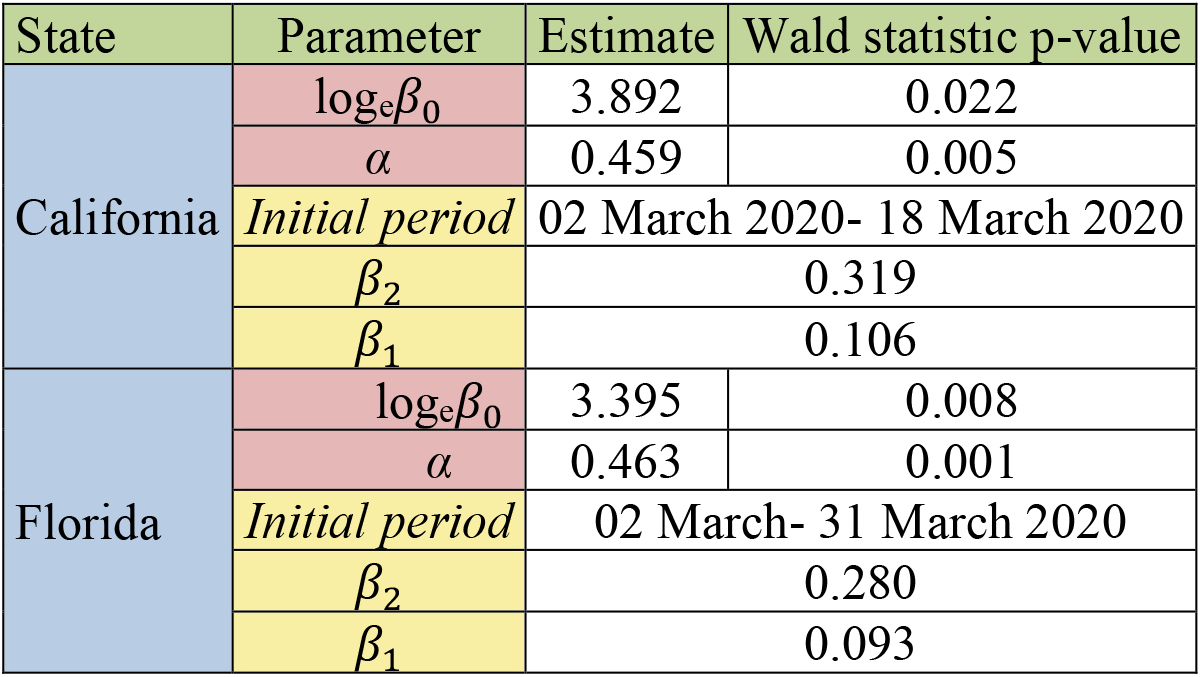
Estimated coefficients of TSIR models with respective p-values of the Wald Chi-square statistic, and resultant estimates of *β*_2_ and *β*_1_.

Using the estimates of CFR and IFR, and the average duration from onset of symptom to death, as reported by Verity *et al*. (2020), we get the estimates of *d*_*1*_ and *d*_*2*_ as follows.

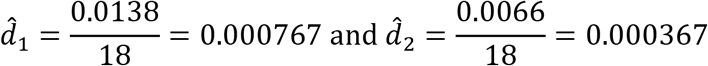

Also, the estimate of recovery rate is 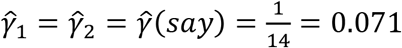. These estimates remain same for both states. So, the initial estimates of average reproduction numbers, *R*_*1*_ and *R*_*2*_, will be given as,

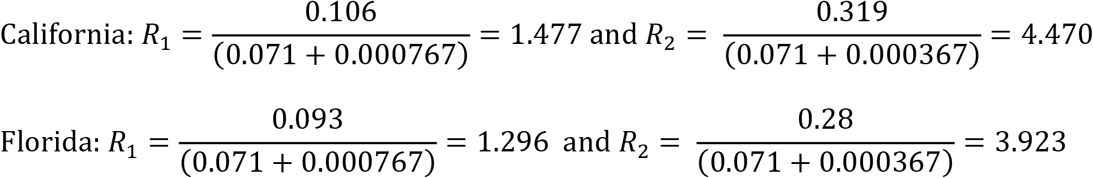

These estimates of *γ, R*_*1*_ and *R*_*2*_ are used to obtain informed hyperparameters of their prior distributions defined in equations (14) and (15). To decide on the hyperparameters of the Beta prior distribution of the time-varying proportion of quarantined infecteds, *p*_*t*,,_ we use the descriptive statistics of its estimates over the observed period. Till the observed time period, the Beta prior distribution of *p*_*t*_ is assumed to have mean equal to 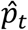, the estimate of *p*_*t*_ based on the calibrated data, and variance equal to the overall variance of the estimates. It is observed that after initial period of around one month, the values of 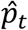 tend to first increase and then settle around some central value. So, for forecasting beyond the observed time period, sample mean and sample variance of the estimates corresponding to the last ten days of the observed period are taken as the mean and variance of the Beta prior distribution of *p*_*t*_. Complete list of prior distributions, along with the values of hyperparameters, used for fitting the state-space SI(Q/F)RD models for the two states are listed below.

California:

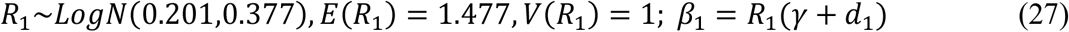

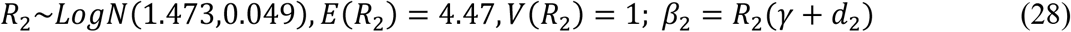

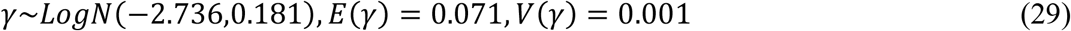

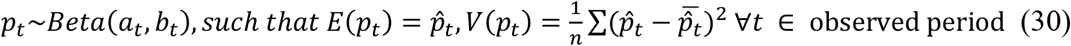

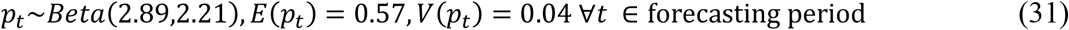

Florida:

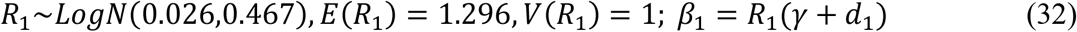

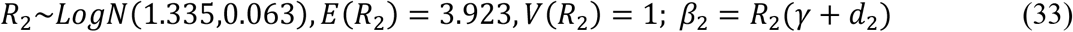

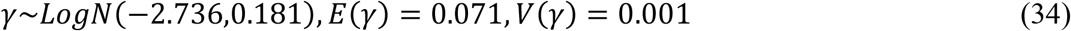

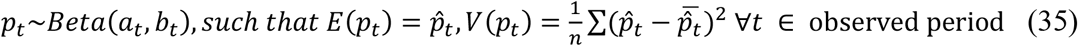

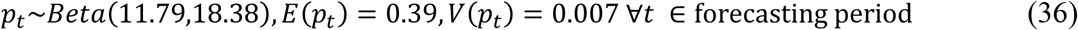

Further, for models pertaining to both states, we assume,

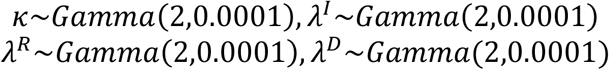

### 3.4. Posterior estimates and forecasts from the state-space SI(Q/F)RD model (results for base case/ intervention)

The Dirichlet-Beta state-space SI(Q/F)RD model defined in section 2.4 is fitted on the calibrated data, using the parameters and hyperparameters obtained in section 3.3. The model is implemented in JAGS platform using R2jags package. Three parallel markov chains were run, each with 20,000 iterations of which first 10,000 were discarded. After thinning at an interval of 10, 1000 posterior simulations were saved from each chain, *i*.*e*., total 3000 posterior simulations were saved for each parameter. We have developed self-written R programming codes for these computations. Posterior estimates of time-invariant parameters along with their standard deviations and 95% credible intervals for the two states are presented in Table 4 and Table 5. Plots of predicted values of the observed process on the number of infecteds 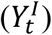 and the number of deaths 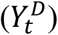, corresponding region of 95% credible intervals, and observed (calibrated) counts till the observed time, are exhibited for the two states in Figures 3 and 4. These results pertain to the base intervention, *i*.*e*., they are predictions based on the assumption that current level of testing (targeted testing) will be carried out throughout the course of the epidemic.

**Table 4:** Posterior estimates of time-invariant parameters of the state-space SI(Q/F)RD model, along with their standard deviations and 95% credible intervals-California.

| Parameter | Posterior mean | Posterior standard deviation | 95% credible interval |
| --- | --- | --- | --- |
| $R_1$ | 0.497 | 0.262 | [0.068, 1.004] |
| $R_2$ | 1.464 | 0.155 | [1.214, 1.813] |
| $\gamma$ | 0.069 | 0.006 | [0.056, 0.081] |
| $\kappa$ | 336063.593 | 47259.956 | [243264.879, 431918.329] |
| $\lambda^D$ | 1355.195 | 718.277 | [397.588, 2632.883] |
| $\lambda^I$ | 1012524.750 | 734717.729 | [1349.955, 2006982.462] |
| $\lambda^R$ | 1633152.503 | 334437.988 | [1073803.103, 2360304.964] |
| $\hat{\beta}_1 = \hat{R}_1(\hat{\gamma} + d_1)$ | 0.035 | | |
| $\hat{\beta}_2 = \hat{R}_2(\hat{\gamma} + d_2)$ | 0.102 | | |

**Table 5:**
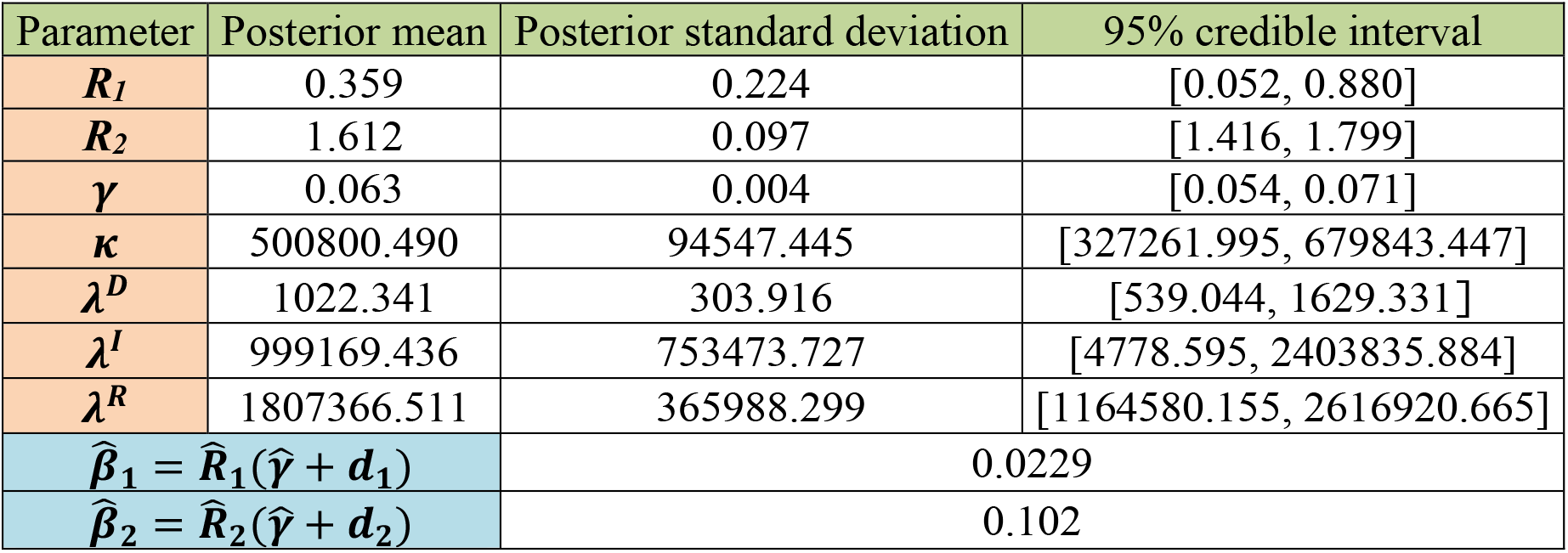
Posterior estimates of time-invariant parameters of the state-space SI(Q/F)RD model, along with their standard deviations and 95% credible intervals-Florida.

| Parameter | Posterior mean | Posterior standard deviation | 95% credible interval |
| --- | --- | --- | --- |
| $R_1$ | 0.359 | 0.224 | [0.052, 0.880] |
| $R_2$ | 1.612 | 0.097 | [1.416, 1.799] |
| $\gamma$ | 0.063 | 0.004 | [0.054, 0.071] |
| $\kappa$ | 500800.490 | 94547.445 | [327261.995, 679843.447] |
| $\lambda^D$ | 1022.341 | 303.916 | [539.044, 1629.331] |
| $\lambda^I$ | 999169.436 | 753473.727 | [4778.595, 2403835.884] |
| $\lambda^R$ | 1807366.511 | 365988.299 | [1164580.155, 2616920.665] |
| $\hat{\beta}_1 = \hat{R}_1(\hat{\gamma} + d_1)$ | 0.0229 | | |
| $\hat{\beta}_2 = \hat{R}_2(\hat{\gamma} + d_2)$ | 0.102 | | |

**Figure 3:**
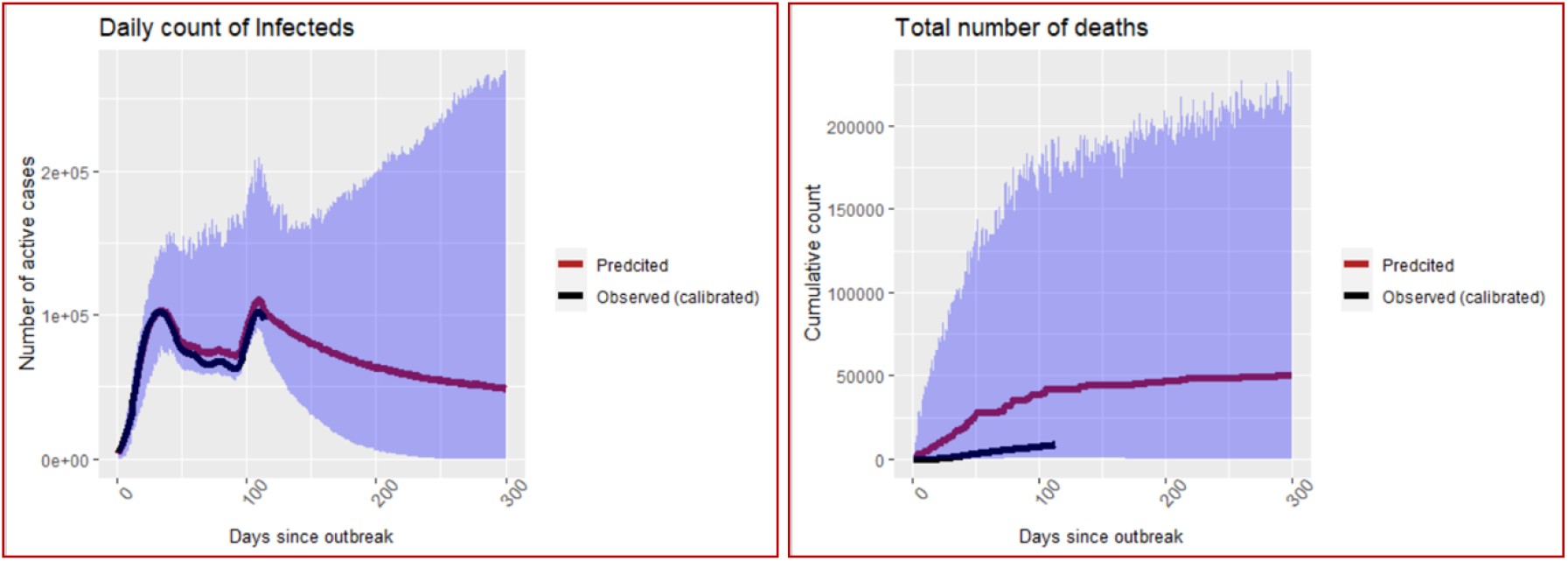
Predictions of number of infecteds and number of deaths in California under the base case/ intervention of targeted testing. The blue shaded ribbon is the region of 95% credible intervals.

**Figure 4:**
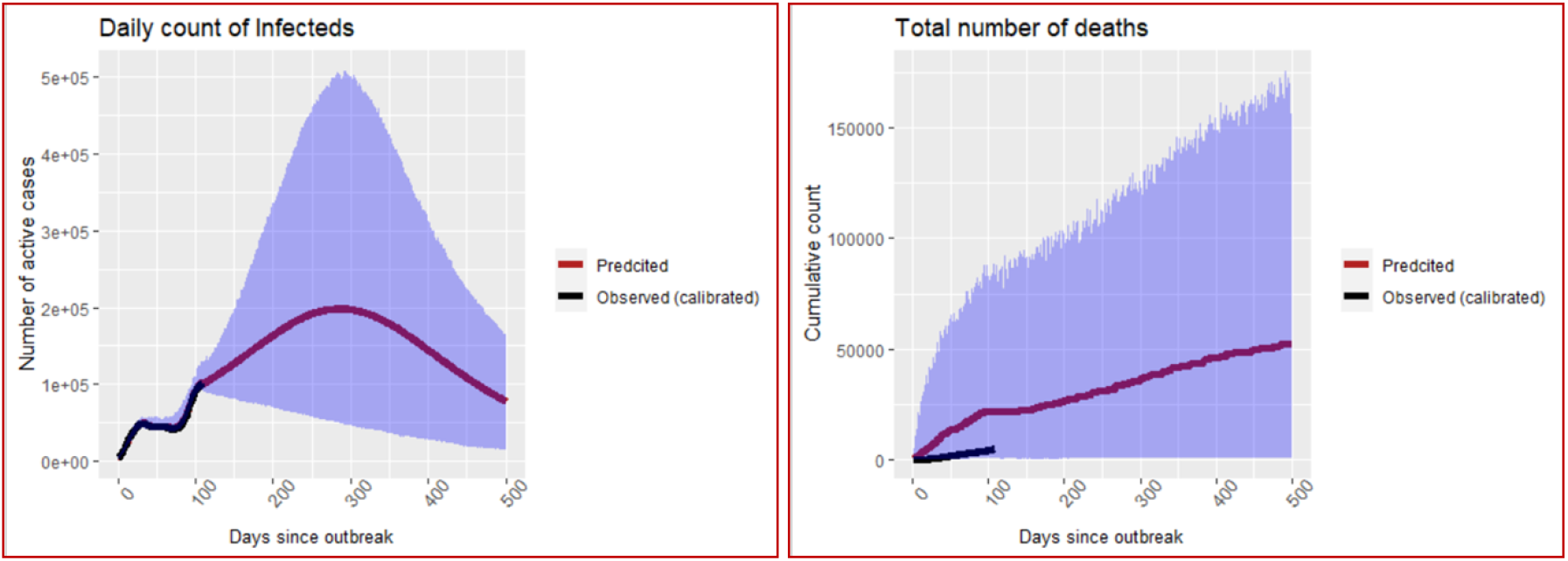
Predictions of number of infecteds and number of deaths in Florida under the base case/ intervention of targeted testing. The blue shaded ribbon is the region of 95% credible intervals.

### 3.5. Predictions under the assumption of extensive random testing (recommended intervention)

Once the posterior estimates of the transmission parameters are obtained from the state-space SI(Q/F)RD model, predictions of observed process under the assumption of extensive random testing are carried out using the steps outlined in section 2.7. Based on the posterior mean and standard deviation of the parameters of the state-space model, following specifications are used for conducting the required simulations to predict the transmission dynamics of the epidemic.

California:

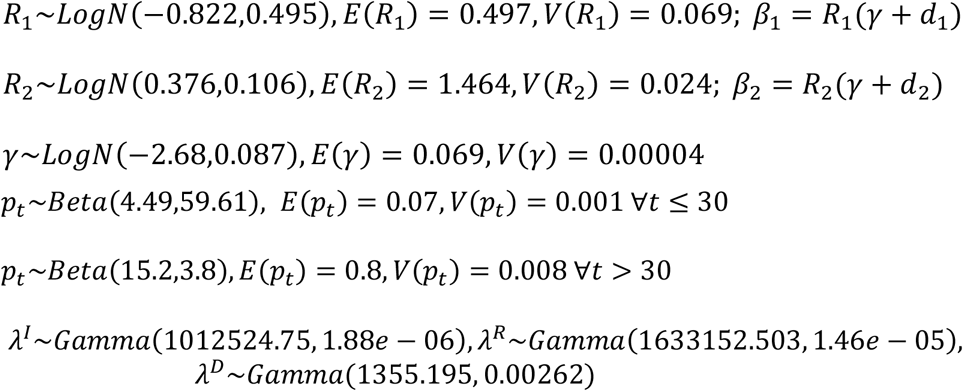

Florida:

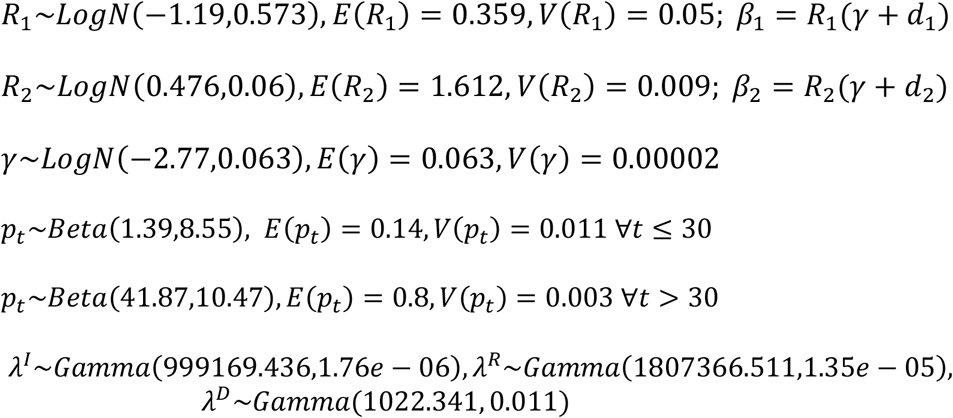

The entire simulation exercise for this section is implemented in R programming through self-written codes. Plots of predicted values of daily number of active infected cases and cumulative deaths, along with their 95% confidence intervals, are shown in Figures 5 and 6. For a comparative assessment of the predictions of transmission trajectory of the epidemic under the two interventions, daily counts of active infecteds and cumulative number of deaths for both cases are plotted together in Figures 7 and 8.

**Figure 5:**
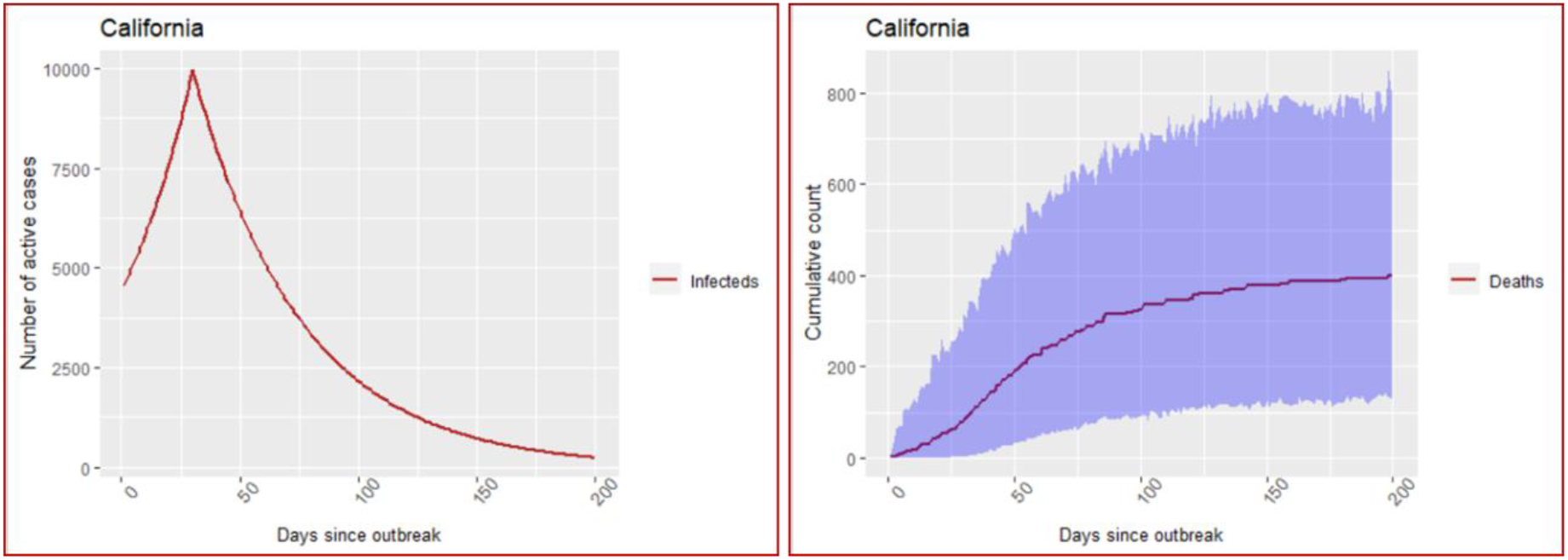
California- Predictions under the assumption of extensive random testing. The blue shaded region depicts the region of 95% confidence intervals based on simulated values. The confidence region for number of infecteds is too narrow to be visible in the graph.

**Figure 6:**
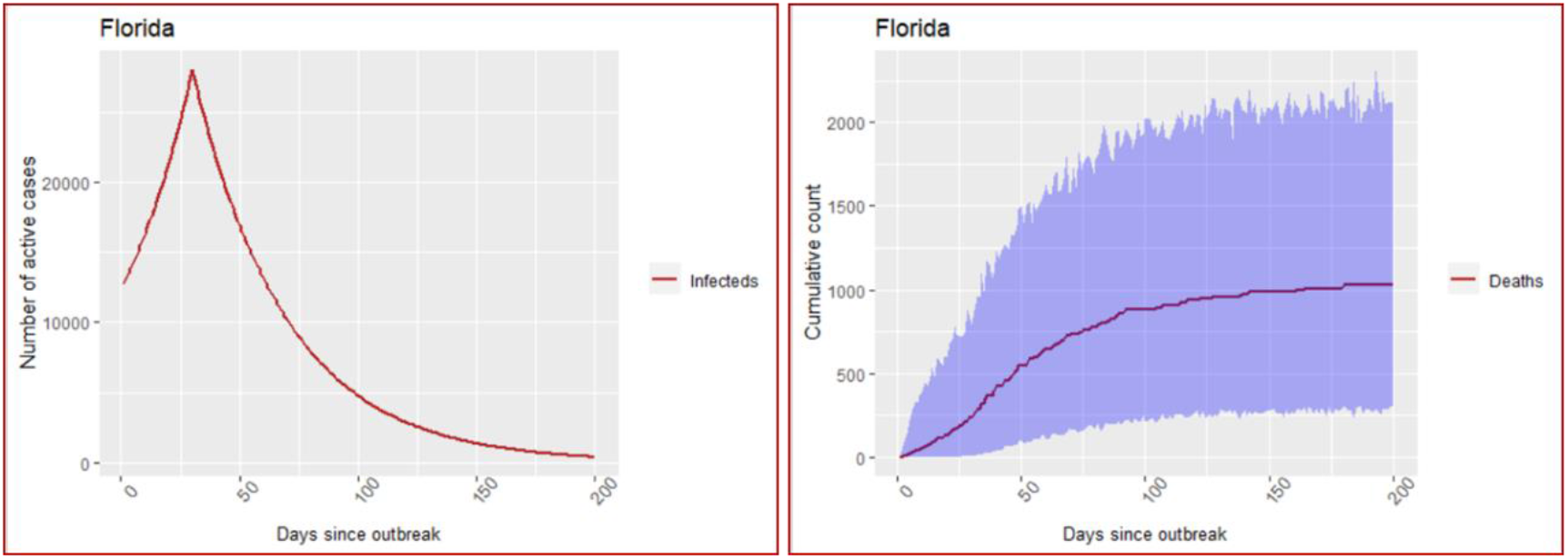
Florida- Predictions under the assumption of extensive random testing. The blue shaded region depicts the region of 95% confidence intervals based on simulated values. The confidence region for number of infecteds is too narrow to be visible in the graph.

**Figure 7:**
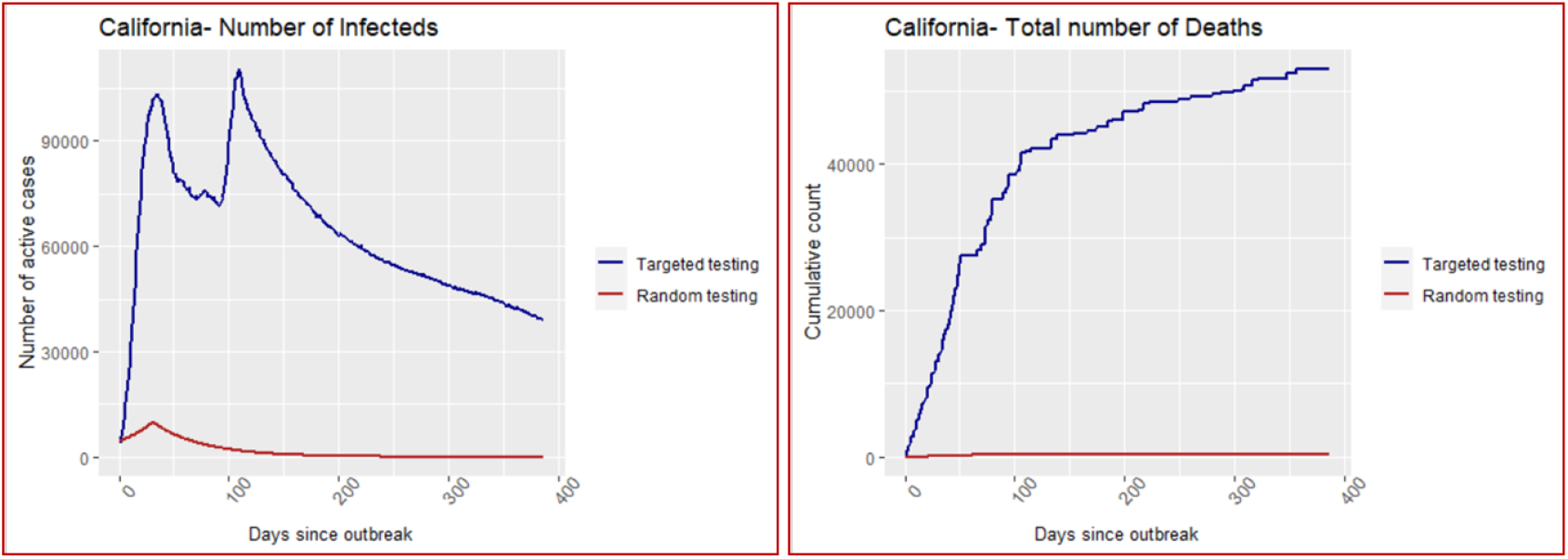
California- Comparative graphs of predictions of cases under both interventions.

**Figure 8:**
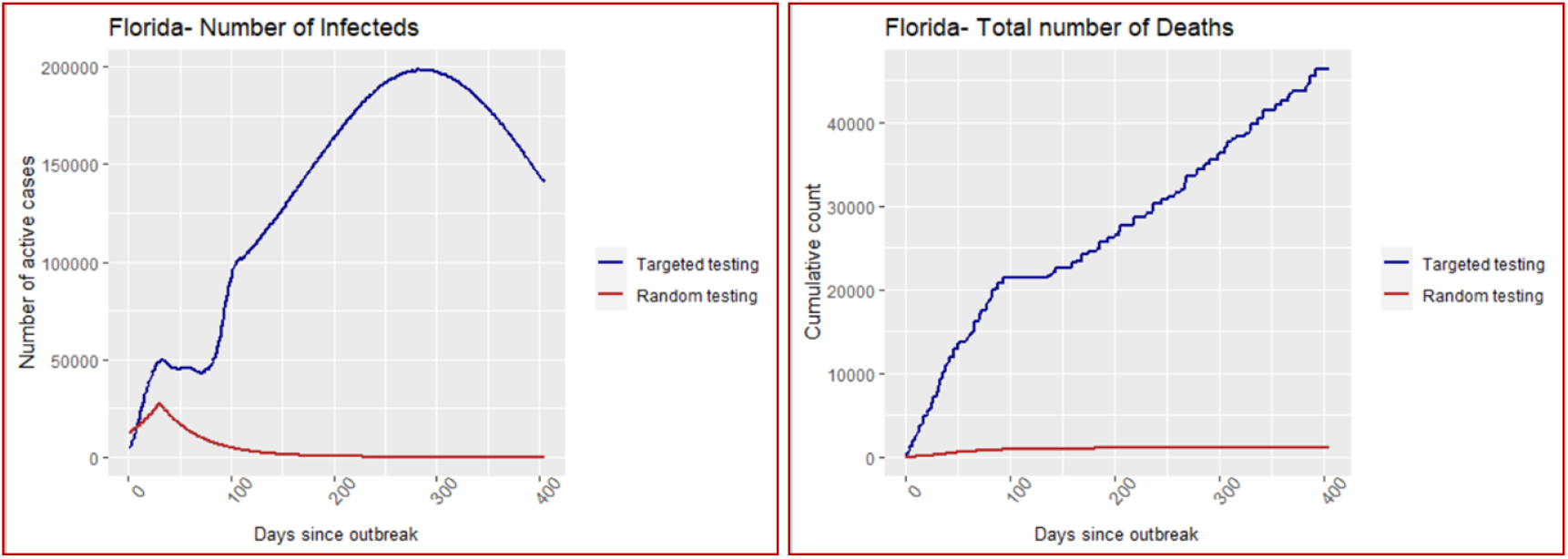
Florida- Comparative graphs of predictions of cases under both interventions.

### 3.6. CEA of extensive random testing over targeted testing

To estimate the cost incremental we first need the estimates of total number of tests to be conducted under both interventions. The rates of positivity of COVID-19 testing, as reported till 29 July 2020, were 7.47% in California and 18.96% in Florida. These percentages were taken as *r*_*1*_ for estimating number of tests under the base intervention of targeted testing. The rates of positivity of tests under the assumption of extensive random testing, *r*_*2*_ are obtained as 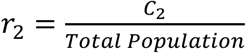 for the two states and are provided in Table 6.

**Table 6:**
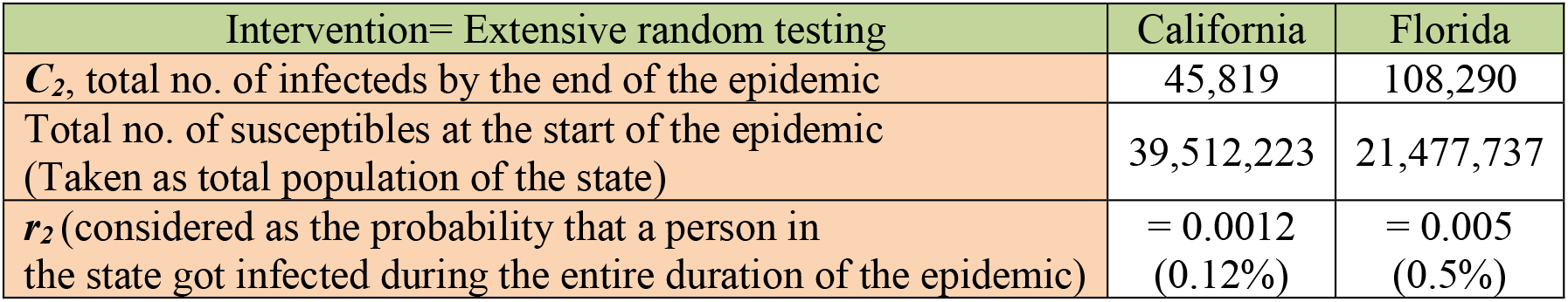
Estimates of rates of positivity of tests under extensive random testing.

| Intervention= Extensive random testing | California | Florida |
| --- | --- | --- |
| $C_2$, total no. of infecteds by the end of the epidemic | 45,819 | 108,290 |
| Total no. of susceptibles at the start of the epidemic<br>(Taken as total population of the state) | 39,512,223 | 21,477,737 |
| $r_2$ (considered as the probability that a person in the state got infected during the entire duration of the epidemic) | = 0.0012<br>(0.12%) | = 0.005<br>(0.5%) |

Values of cost incremental, changes in outcome measures (both in terms of number of infections and number of deaths), and ICERs are calculated by implementing these values of *r*_*1*_ and *r*_*2*_ in the steps discussed in part e. of section 2.7. Cost of COVID-19 test (RT-PCR) varies considerably across USA. However, leaving out some extreme cases, the average cost per unit of the RT-PCR test is around $100 in USA [Kliff (2020)]. We have used this average cost for evaluating cost incremental owing to increment in the number of tests. Results on the difference in number of tests, cost increment (or decrement), and changes in the outcomes of number of infections and number of deaths, on using the proposed intervention ‘extensive random testing’ over the base intervention ‘targeted testing’, are furnished in Table 7.

**Table 7:** Changes in outcomes and costs on using extensive random testing instead of targeted testing as the intervention to contain the spread of SARS-CoV-2 infections.

| State | Total Confirmed Cases | Total Detected Cases | Number of Tests | Number of Deaths |
| --- | --- | --- | --- | --- |
| Predictions under the base case (Base intervention = Targeted testing) |  |  |  |  |
| California | 2384143 | 1328158 | 17779893 | 58292 |
| Florida | 4793903 | 1873747 | 9882632 | 58937 |
| Predictions under the ideal case (Recommended intervention = Extensive random testing)<br>(Case A- Number of tests estimated using $r_2$) | | | | |
| California | 45819 | 41237 | 35560914 | 405 |
| Florida | 108290 | 97461 | 19329963 | 1039 |
| Predictions under the ideal case (Recommended intervention = Extensive random testing)<br>(Case B- Assuming that everyone was tested by the end of the epidemic) |  |  |  |  |
| California | 45819 | 41237 | 39512223 | 405 |
| Florida | 108290 | 97461 | 21477737 | 1039 |
| Changes in Cost and Outcomes |  |  |  |  |
| State | Reduction in occurrence of infection | Reduction in occurrence of death | Tests incremental | Testing cost incremental (\$) |
| Case A-Cost-effectiveness of extensive random testing with respect to targeted testing |  |  |  |  |
| California | 2338324 | 57887 | 17781021 | 1778102100 |
| Florida | 4685613 | 57898 | 9447331 | 944733100 |
| Case B-Cost-effectiveness of extensive random testing with respect to targeted testing |  |  |  |  |
| California | 2338324 | 57887 | 21732330 | 2173233000 |
| Florida | 4685613 | 57898 | 11595105 | 1159510500 |

## 4. Discussion

Estimates of the proportion of reported cases, *p*_*t*_, based on the calibrated data give us a good idea about the seriousness of the problem of underreporting. As can be seen from the summary provided in Table 2, the first 30 days of the outbreak of the COVID-19 epidemic have experienced extremely high level of underreporting, with the average percentages of underreporting for the two states being 96.6% (California) and 93.9% (Florida). This can be attributed to the lack of appropriate testing facilities, including resources like testing kits, and absence of proper response from the government (or lack of seriousness and foresightedness on the part of the policy makers) during the initial stage of the pandemic. With time, the proportion of reported cases increases steadily, and after a couple of months, it tends to converge around the average values of 57% for California and 39% for Florida. These average values of *p*_*t*_, calculated at the later stage of the pandemic, are representative of the nature and capacity of testing policy of the states. For the same reason, the hyperparameters of the prior distributions of *p*_*t*_ for the purpose of forecasting are based on these later-stage averages. That is, due to the lack of extensive random testing in these states, around 43% infected cases in California and 61% infected cases in Florida, on an average, go unreported. These percentages of infecteds are undetected and are not quarantined, and they remain infectious for a much longer period than their quarantined counterparts, moving freely among the susceptibles. The SI(Q/F)RD epidemic model proposed by us is based on this hypothesis and the hypothesis is strongly supported by the posterior estimates of the transmission parameters obtained from the Dirichlet-Beta state-space SI(Q/F)RD model. Posterior estimates of average reproduction numbers associated with quarantined infecteds are 0.497 (sd: 0.262) and 0.359 (sd: 0.224), and for the undetected infecteds are 1.464 (sd: 0.155) and 1.612 (sd: 0.097) for California and Florida respectively. This clearly indicates that if almost all infecteds were quarantined; the number of active cases would have declined sharply bringing the epidemic to an end at a very early stage. However, quarantining almost all infecteds, in the presence of a large proportion of asymptomatic cases, requires extensive amount of daily testing. This is clearly missing in both states under consideration, as indicated by very high rates of positivity of tests and high CFR values based on reported data for the two states. So, considering extensive random testing as an intervention to contain the epidemic is very essential for understanding its effectiveness both in terms of outcome (reducing infections and deaths) and cost (extra cost required for extensive testing instead of targeted testing).

For both states, CEA of extensive random testing over targeted testing has yielded very strong results in favour of the former; refer Table 7 and Table 8. Citing uncertainties because of some unknown factors and leaving some space for errors in testing, even if we assume that 80% of the infecteds can be detected and quarantined using extensive random testing, a total of around 2.3 million people in California and 4.7 million people in Florida could be saved from the infection by the end of the epidemic if extensive random testing was used instead of targeted testing. Further, it is estimated that around 58 thousand deaths due to COVID-19 could be averted in each state if the states resorted to extensive random testing (after first month of outbreak) instead of targeted testing. These are huge expected gains for humanity, especially when every single life matter for us. The ICER values (in terms of number of tests) suggest that, on an average, only around 9 and 2 additional number of tests would be required in total to save one extra person from getting infected in California and Florida, respectively, by the time the epidemic ends. That is, around 760-929 USD (California) and 202-247 USD (Florida) additional expenditure on COVID-19 tests would be required to save every additional person from getting infected. Number of additional tests required to save one additional death from COVID-19 is estimated to be around 307-375 for California and 163-200 for Florida. That is, on using extensive random testing over targeted testing, one extra loss of life due to COVID-19 can be averted on an additional expenditure of around 30717-37543 USD in California and around 16317-20027 USD in Florida.

**Table 8:** Incremental cost effectiveness ratios associated with extensive random testing as compared to targeted testing.

| Incremental Cost Effectiveness Ratio- ICER |  |  |
| --- | --- | --- |
| State | Number of extra tests per unit reduction in infection |  |
|  | Case A* | Case B** |
| California | 8 | 9 |
| Florida | 2 | 2 |
|  | Number of extra tests per unit reduction in death |  |
|  | Case A | Case B |
| California | 307 | 375 |
| Florida | 163 | 200 |
| ICER<br>(infections) | Additional testing cost per unit reduction in infection (\$) | |
|  | Case A | Case B |
| California | 760 | 929 |
| Florida | 202 | 247 |
| ICER<br>(deaths) | Additional testing cost per unit reduction in death (\$) | |
|  | Case A | Case B |
| California | 30717 | 37543 |
| Florida | 16317 | 20027 |

\*Case A- Number of tests estimated using $r_2$ .
\*\*Case B- Assuming that everyone was tested by the end of the epidemic.

### Comparing forecasted deaths with post-study published estimate of excess deaths-comments on the validity of the assumptions underlining the proposed state-space SI(Q/F)RD model

Since the true number of infecteds, detected plus undetected, remain latent in the population, it is not possible to compare forecasted values with the true values. However, comparing cumulative number of deaths forecasted by the state-space SI(Q/F)RD model with the estimated values of excess deaths can serve as a potent alternative to assess predictive efficiency of the model. Estimates of epidemiological parameters and predictions obtained from the state-space SI(Q/F)RD model are based on the daily time-series data on number of cases reported till 11 July 2020 and the weekly estimates of excess deaths available till the same date. Predictive accuracy of the model will be determined by its ability to forecast true values beyond the training period of the model. From this perspective, we have plotted the forecasted time-series of cumulative number of deaths obtained from the fitted SI(Q/F)RD model along with the weekly estimated excess deaths due to COVID-19 till 14 November 2020. The estimates of excess deaths due to COVID-19 have been retrieved from the website of CDC (https://www.cdc.gov/nchs/nvss/vsrr/covid19/excess_deaths.html) on 4 December 2020. Striking difference in the estimated values of average reproduction numbers associated with detected (quarantined) cases and undetected cases suggest that the assumption regarding future values of proportion of detected cases plays a crucial role in ascertaining high predictive accuracy of the model. In other words, accuracy of the predictions from the proposed SI(Q/F)RD model relies greatly on the validity of the assumption regarding future testing policy in the region-*i*.*e*., on whether the testing capacity relative to the number of true cases is expected to increase, remain unchanged or decrease over time. To stress upon this argument, observed time-series of rates of positivity of COVID-19 tests are also plotted alongside the comparative plots of the forecasted number of deaths. Figure 9 and Figure 10 present these plots for California and Florida, respectively. In both of these figures, the left panel shows the comparative trends of cumulative number of deaths (predicted and excess) and the right panel presents the rate of positive tests over time. The trend line in blue in the right panel shows the average percentage of tests that were positive over the last seven days, *i*.*e*., a seven-day moving average of percentage of positive tests. The time-series plots of rates of positive tests for the two states are sourced from the website of Johns Hopkins University on 08 December 2020 [https://coronavirus.jhu.edu/testing/testing-positivity].

**Figure 9:**
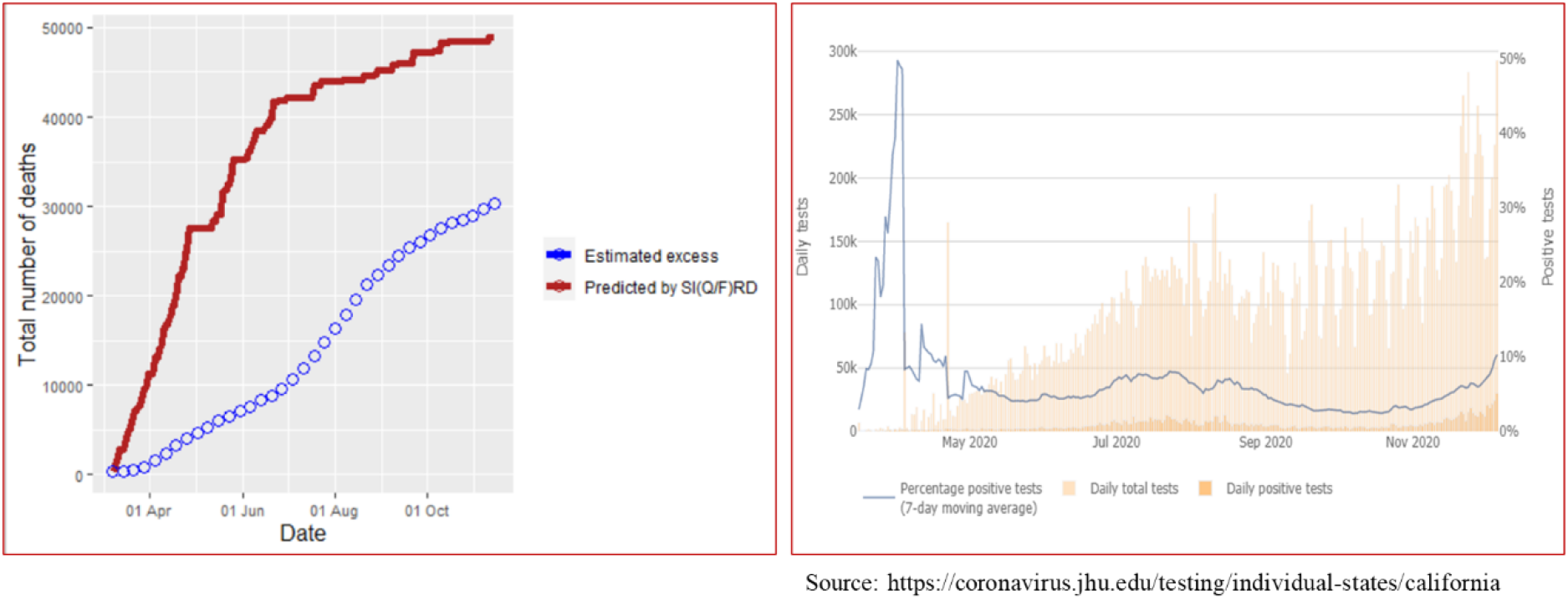
California- Left panel shows comparison of cumulative number of deaths predicted by the SI(Q/F)RD model with the estimates excess deaths due to COVID-19. Right panel shows trend line of seven-day moving average of percentage of positive tests, along with daily total number of tests and daily total number of positive tests.

**Figure 10:**
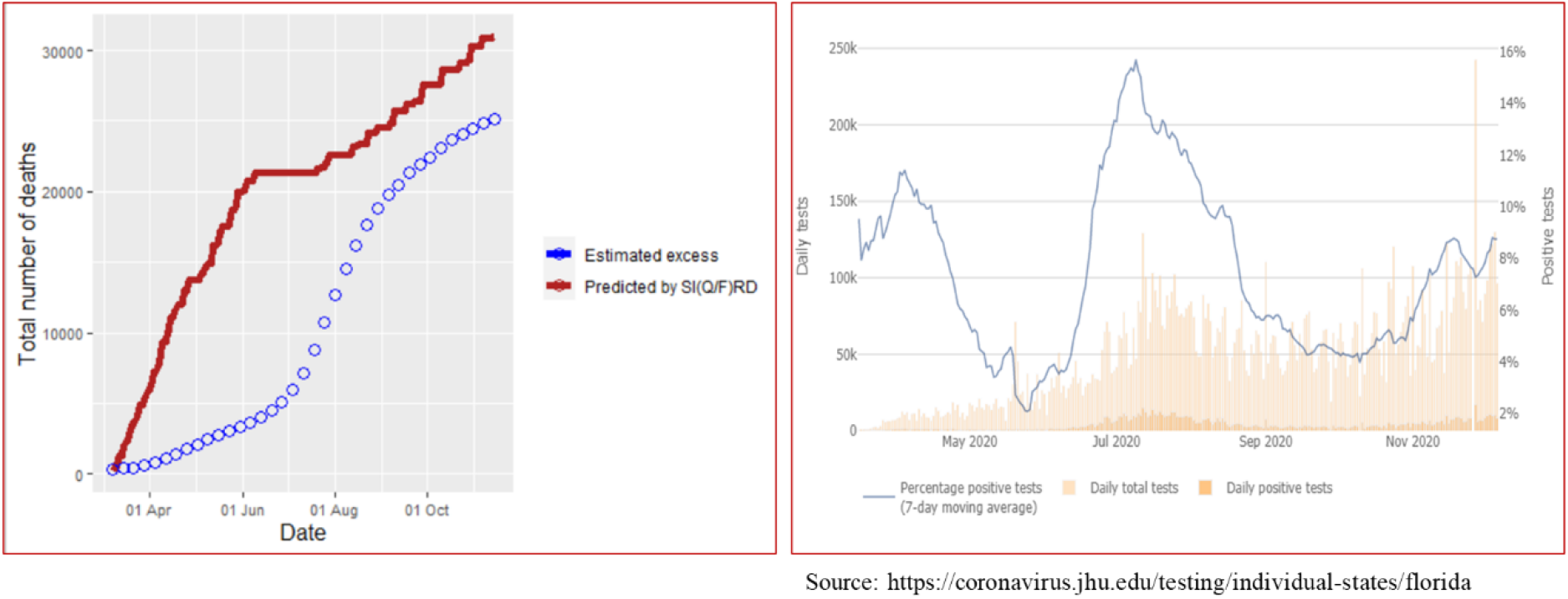
Florida- Left panel shows comparison of cumulative number of deaths predicted by the SI(Q/F)RD model with the estimates excess deaths due to COVID-19. Right panel shows trend line of seven-day moving average of percentage of positive tests, along with daily total number of tests and daily total number of positive tests.

In the case of California, there is a considerable difference between the predicted number of deaths and estimated excess deaths due to COVID-19. However, the difference is majorly in the scale of the values, and the two trend lines look similar in shape over time. Possible reason for the difference in the scale can be explained by analysing the trend line of rate of positive tests. The percentage of positive tests in California was extremely high during March-April, but although it started dropping exceptionally towards the end of April, it remained around 10% till July. However, September onwards the rate of positivity came down below 5%, the WHO recommended threshold. The steep rise in total number of tests performed daily, as shown by the pink towers in the graph, clearly explains this change. That is, California experienced a drastic change in testing capacity in the period of forecasting. Since the hyperparameters of the model corresponding to the proportion of detected cases were defined based on the state of rate of positivity till July, the model tends to give overestimated forecasts of total number of deaths. Increasing the proportion of detected cases in the model as per the increase in testing capacity of the region would result in decrease in the total number of deaths. This is because the estimated rate of transmission for detected (quarantined) cases is relatively much lower than that of the undetected cases.

The scenario of rate of positive tests over time looks entirely different for Florida. Percentage of positive tests dipped below 5% for only two brief periods and it remained high for most of the time. That is, except for few short periods, the testing capacity has remained below par. Insufficient testing is also indicated by the fact that the trend line of the rate of positive tests is mostly parallel to the changes in the peaks of the total number of tests conducted per day. That is, it suggests that increase in the number of tests was not sufficient to reduce the rate of positive tests. Ideally, in the presence of sufficient amount of random testing, rate of positive tests should decrease with increase in the number of tests- as can be seen in the case of California. In other words, no significant change in the testing policy of Florida is observed in the forecasting period. This further implies that the hyperparameters defined for the proportion of detected cases in the state-space SI(Q/F)RD model remained valid for the forecasting period. Consequently, the forecasted values of cumulative number of deaths are much closer to the estimates of excess deaths in the case of Florida as compared to that of California. These results reaffirm the inevitable impact of testing capacity on the transmission dynamics of the pandemic, which forms the conceptual backbone of the proposed state-space SI(Q/F)RD model.

## 5. Conclusion

We have provided a detailed framework of data calibration and flexible epidemic modelling for conducting CEA of non-medical interventions for containing epidemics like COVID-19. The structure of the proposed SI(Q/F)RD model allows for adjusting the trajectory of the epidemic in terms of level of underreporting. The Dirichlet-Beta state-space formulation of the SI(Q/F)RD model provides a flexible approach to the estimation and prediction of both time-invariant and time-varying transmission parameters of the epidemic. The state-space model allows for uncertainties in the transmission dynamics over time and thus, it can be considered to be superior to its deterministic counterpart. The proposed method, based on TSIR, for estimating hyperparameters of prior distributions of transmission rates (or reproduction rates) is aimed at improving the posterior estimates by enriching the state-space model with strong prior information.

The results of CEA conclude that extensive random testing, which has been strongly recommended by WHO, is significantly cost-effective over targeted testing. Since the *R*_*0*_ values associated with quarantined infecteds in both states are estimated to be below 1, extensive random testing, resulting in quarantining of at least 80% infecteds, is expected to result in the epidemic to end quite quickly as compared to the case of targeted testing. So, targeted testing may imply a smaller number of tests over a much longer period of time, while extensive testing means a very high number of tests for a much shorter period of time. This simple logic is corroborated by the ICER values obtained from the CEA of extensive random testing over targeted testing. For California, if the state is willing to conduct around 9 extra tests (or spend around 900 USD extra amount on testing) for saving one additional person from getting infected, or if the state is willing to conduct around 375 extra tests (or spend around 37500 USD extra amount on testing) for saving one additional person from dying due to COVID-19, extensive random testing can be considered as cost-effective over targeted testing. While for Florida, willingness to spend an extra amount of around 200 USD (2 extra tests) for saving one additional person from getting infected, or willingness to spend an extra amount of around 20,000 USD (200 extra tests) for saving one additional person from dying due to COVID-19, renders extensive random testing as cost-effective over targeted testing.

### Limitations and further scope of research

In the state-space SI(Q/F)RD model, we have taken the rate of death as a fixed (known) parameter. Using posterior estimate of the death rate, instead of defining it as a fixed parameter, may further improve the overall predictions from the model by introducing stochastic uncertainty. Also, time varying transmission rates can be introduced in the model using the modifier functions described in Deo *et al*. (2020) to make the model more robust.

## Data Availability

Data has been procured from the github repository of Johns Hopkins University and the website of the CDC, USA.

https://github.com/CSSEGISandData/COVID-19

https://www.cdc.gov/nchs/nvss/vsrr/covid19/excess_deaths.html

## Funding

This research did not receive any specific grant from funding agencies in the public, commercial, or not-for-profit sectors.

## Appendix-A

**Table A.1:** Calibrated data of California and Florida.

| Date | I_cali | p_obs_cali | R_cali | D_cali | I_florida | p_obs_florida | R_florida | D_florida |
| --- | --- | --- | --- | --- | --- | --- | --- | --- |
| 2-Mar-20 | 1449 | 0.0062 | 0 | 0 | 217 | 0.0046 | 0 | 0 |
| 3-Mar-20 | 2963 | 0.0025 | 80 | 0 | 569 | 0.0028 | 10 | 0 |
| 4-Mar-20 | 4575 | 0.0055 | 279 | 1 | 1115 | 0.0000 | 44 | 0 |
| 5-Mar-20 | 6337 | 0.0076 | 618 | 1 | 1785 | 0.0014 | 99 | 0 |
| 6-Mar-20 | 8300 | 0.0033 | 1046 | 1 | 2604 | 0.0000 | 222 | 0 |
| 7-Mar-20 | 10497 | 0.0080 | 1603 | 1 | 3565 | 0.0037 | 348 | 0 |
| 8-Mar-20 | 12817 | 0.0045 | 2399 | 1 | 4573 | 0.0024 | 570 | 2 |
| 9-Mar-20 | 15637 | 0.0017 | 3202 | 1 | 5727 | 0.0021 | 865 | 2 |
| 10-Mar-20 | 18592 | 0.0106 | 4304 | 2 | 6976 | 0.0012 | 1283 | 2 |
| 11-Mar-20 | 21862 | 0.0076 | 5525 | 4 | 8373 | 0.0050 | 1698 | 2 |
| 12-Mar-20 | 25183 | 0.0086 | 7204 | 4 | 9956 | 0.0030 | 2144 | 2 |
| 13-Mar-20 | 28787 | 0.0109 | 9107 | 4 | 11429 | 0.0092 | 2845 | 2 |
| 14-Mar-20 | 32367 | 0.0149 | 11606 | 12 | 13140 | 0.0134 | 3447 | 8 |
| 15-Mar-20 | 37606 | 0.0003 | 13024 | 22 | 15458 | 0.0142 | 3661 | 12 |
| 16-Mar-20 | 43209 | 0.0256 | 14583 | 34 | 17781 | 0.0004 | 4016 | 15 |
| 17-Mar-20 | 48952 | 0.0190 | 16361 | 49 | 20035 | 0.0315 | 4586 | 17 |
| 18-Mar-20 | 54635 | 0.0169 | 18415 | 66 | 22286 | 0.0390 | 5304 | 19 |
| 19-Mar-20 | 60181 | 0.0225 | 20748 | 86 | 24405 | 0.0414 | 6225 | 22 |
| 20-Mar-20 | 65496 | 0.0291 | 23454 | 108 | 26514 | 0.0414 | 7299 | 27 |
| 21-Mar-20 | 70606 | 0.0206 | 26507 | 133 | 28550 | 0.0610 | 8516 | 35 |
| 22-Mar-20 | 75324 | 0.0287 | 30093 | 162 | 30523 | 0.0711 | 9939 | 45 |
| 23-Mar-20 | 79730 | 0.0550 | 34060 | 195 | 32345 | 0.0644 | 11582 | 58 |
| 24-Mar-20 | 83739 | 0.0506 | 38491 | 233 | 34097 | 0.0521 | 13366 | 73 |
| 25-Mar-20 | 87259 | 0.0551 | 43406 | 276 | 35714 | 0.0745 | 15355 | 90 |
| 26-Mar-20 | 90316 | 0.1063 | 48777 | 326 | 37266 | 0.1826 | 17479 | 110 |
| 27-Mar-20 | 92797 | 0.0907 | 54718 | 382 | 38727 | 0.1441 | 19763 | 133 |
| 28-Mar-20 | 94737 | 0.0518 | 61267 | 444 | 40056 | 0.2247 | 22250 | 158 |
| 29-Mar-20 | 96359 | 0.0866 | 68272 | 513 | 41370 | 0.1212 | 24894 | 186 |
| 30-Mar-20 | 97665 | 0.1474 | 75658 | 589 | 42624 | 0.3024 | 27668 | 216 |
| 31-Mar-20 | 98899 | 0.1209 | 83253 | 673 | 43860 | 0.3019 | 30603 | 248 |
| 1-Apr-20 | 99983 | 0.1321 | 90990 | 765 | 45097 | 0.0508 | 33606 | 283 |
| 2-Apr-20 | 100788 | 0.1561 | 98927 | 864 | 46213 | 0.4807 | 36728 | 320 |
| 3-Apr-20 | 101418 | 0.1398 | 106961 | 968 | 47172 | 0.3019 | 39933 | 359 |
| 4-Apr-20 | 101761 | 0.0952 | 115134 | 1075 | 47931 | 0.3097 | 43263 | 400 |
| 5-Apr-20 | 101892 | 0.2591 | 123372 | 1184 | 48549 | 0.2006 | 46661 | 442 |
| 6-Apr-20 | 101726 | 0.1188 | 131760 | 1295 | 48994 | 0.2454 | 50158 | 486 |
| 7-Apr-20 | 101444 | 0.1673 | 140117 | 1408 | 49321 | 0.3138 | 53699 | 531 |
| 8-Apr-20 | 100961 | 0.1992 | 148456 | 1523 | 49510 | 0.2377 | 57304 | 578 |
| 9-Apr-20 | 100134 | 0.1276 | 156848 | 1639 | 49521 | 0.2418 | 61013 | 626 |
| 10-Apr-20 | 99040 | 0.1842 | 165289 | 1756 | 49375 | 0.3163 | 64806 | 675 |
| 11-Apr-20 | 97575 | 0.0892 | 173811 | 1873 | 49005 | 0.2664 | 68749 | 725 |
| 12-Apr-20 | 95891 | 0.1546 | 182407 | 1990 | 48586 | 0.3872 | 72740 | 776 |
| 13-Apr-20 | 93912 | 0.1659 | 191153 | 2107 | 47958 | 0.3171 | 76867 | 828 |
| 14-Apr-20 | 91738 | 0.2182 | 199876 | 2225 | 47240 | 0.1721 | 81083 | 881 |
| 15-Apr-20 | 89508 | 0.1989 | 208508 | 2345 | 46617 | 0.2445 | 85274 | 936 |
| 16-Apr-20 | 87084 | 0.1554 | 217188 | 2466 | 46076 | 0.2310 | 89382 | 992 |
| 17-Apr-20 | 84806 | 0.2348 | 225647 | 2589 | 45608 | 0.3925 | 93415 | 1050 |
| 18-Apr-20 | 82661 | 0.2148 | 233973 | 2712 | 45212 | 0.2034 | 97375 | 1109 |
| 19-Apr-20 | 80747 | 0.1443 | 242142 | 2834 | 44843 | 0.2277 | 101308 | 1168 |
| 20-Apr-20 | 78971 | 0.3497 | 250246 | 2955 | 44550 | 0.2065 | 105166 | 1226 |
| 21-Apr-20 | 77486 | 0.2782 | 258061 | 3074 | 44387 | 0.2200 | 108968 | 1283 |
| 22-Apr-20 | 76352 | 0.2870 | 265600 | 3191 | 44347 | 0.1201 | 112648 | 1339 |
| 23-Apr-20 | 75310 | 0.3425 | 272976 | 3306 | 44346 | 0.3623 | 116290 | 1394 |
| 24-Apr-20 | 74567 | 0.2782 | 280055 | 3419 | 44416 | 0.2394 | 119862 | 1448 |
| 25-Apr-20 | 73975 | 0.1571 | 286986 | 3529 | 44515 | 0.0828 | 123406 | 1501 |
| 26-Apr-20 | 73507 | 0.1868 | 293725 | 3635 | 44690 | 0.1878 | 126875 | 1553 |
| 27-Apr-20 | 73263 | 0.2216 | 300243 | 3738 | 44826 | 0.1642 | 130384 | 1604 |
| 28-Apr-20 | 73103 | 0.1891 | 306608 | 3837 | 44889 | 0.1924 | 133967 | 1654 |
| 29-Apr-20 | 72922 | 0.4194 | 312851 | 3934 | 44950 | 0.0944 | 137552 | 1704 |
| 30-Apr-20 | 72685 | 0.2300 | 319007 | 4029 | 45022 | 0.1364 | 141127 | 1753 |
| 1-May-20 | 72232 | 0.3230 | 325238 | 4121 | 45119 | 0.2827 | 144677 | 1802 |
| 2-May-20 | 71641 | 0.2307 | 331464 | 4211 | 45201 | 0.2010 | 148241 | 1852 |
| 3-May-20 | 70847 | 0.2753 | 337822 | 4299 | 45305 | 0.1688 | 151783 | 1902 |
| 4-May-20 | 70073 | 0.1736 | 344161 | 4386 | 45224 | 0.2294 | 155437 | 1952 |
| 5-May-20 | 69274 | 0.4551 | 350525 | 4473 | 45120 | 0.1560 | 159042 | 2002 |
| 6-May-20 | 68479 | 0.3822 | 356884 | 4561 | 44915 | 0.1668 | 162675 | 2052 |
| 7-May-20 | 67732 | 0.2676 | 363267 | 4650 | 44626 | 0.2516 | 166320 | 2102 |
| 8-May-20 | 67050 | 0.2886 | 369585 | 4739 | 44336 | 0.1186 | 169965 | 2153 |
| 9-May-20 | 66509 | 0.4782 | 375833 | 4829 | 44040 | 0.2452 | 173616 | 2204 |
| 10-May-20 | 66016 | 0.1751 | 382107 | 4918 | 43772 | 0.1891 | 177239 | 2255 |
| 11-May-20 | 65641 | 0.2910 | 388335 | 5007 | 43427 | 0.1308 | 180866 | 2306 |
| 12-May-20 | 65506 | 0.2745 | 394395 | 5096 | 43081 | 0.2973 | 184494 | 2357 |
| 13-May-20 | 65504 | 0.3023 | 400323 | 5184 | 42752 | 0.1608 | 188105 | 2408 |
| 14-May-20 | 65774 | 0.3406 | 406052 | 5272 | 42473 | 0.2631 | 191739 | 2459 |
| 15-May-20 | 66173 | 0.2993 | 411653 | 5359 | 42178 | 0.2948 | 195389 | 2510 |
| 16-May-20 | 66646 | 0.3338 | 417182 | 5444 | 41961 | 0.2159 | 199033 | 2561 |
| 17-May-20 | 67003 | 0.2396 | 422756 | 5527 | 41882 | 0.2411 | 202612 | 2612 |
| 18-May-20 | 67391 | 0.2248 | 428301 | 5608 | 41990 | 0.2575 | 206076 | 2663 |
| 19-May-20 | 67805 | 0.3735 | 433822 | 5687 | 42240 | 0.1572 | 209471 | 2714 |
| 20-May-20 | 68022 | 0.3793 | 439469 | 5765 | 42530 | 0.1640 | 212826 | 2765 |
| 21-May-20 | 68096 | 0.3516 | 445114 | 5843 | 42850 | 0.3474 | 216152 | 2815 |
| 22-May-20 | 68012 | 0.3937 | 450772 | 5921 | 43158 | 0.2321 | 219491 | 2864 |
| 23-May-20 | 67762 | 0.4091 | 456524 | 5999 | 43452 | 0.2056 | 222845 | 2912 |
| 24-May-20 | 67327 | 0.2733 | 462387 | 6078 | 43880 | 0.2187 | 226138 | 2959 |
| 25-May-20 | 66834 | 0.4360 | 468308 | 6157 | 44476 | 0.2464 | 229408 | 3006 |
| 26-May-20 | 66245 | 0.5560 | 474252 | 6237 | 45269 | 0.1438 | 232698 | 3053 |
| 27-May-20 | 65542 | 0.2945 | 480236 | 6318 | 46133 | 0.1085 | 236062 | 3100 |
| 28-May-20 | 64809 | 0.5114 | 486249 | 6400 | 47244 | 0.1638 | 239398 | 3146 |
| 29-May-20 | 64206 | 0.5198 | 492204 | 6483 | 48481 | 0.2756 | 242825 | 3192 |
| 30-May-20 | 63743 | 0.5914 | 498091 | 6566 | 49851 | 0.2050 | 246337 | 3238 |
| 31-May-20 | 63316 | 0.3685 | 504014 | 6650 | 51376 | 0.1600 | 249910 | 3285 |
| 1-Jun-20 | 63010 | 0.4859 | 509961 | 6734 | 53119 | 0.1382 | 253555 | 3332 |
| 2-Jun-20 | 62889 | 0.4282 | 515795 | 6818 | 55189 | 0.1224 | 257162 | 3380 |
| 3-Jun-20 | 63160 | 0.3590 | 521455 | 6901 | 57441 | 0.2363 | 260803 | 3429 |
| 4-Jun-20 | 63862 | 0.4473 | 526974 | 6984 | 59937 | 0.2448 | 264416 | 3479 |
| 5-Jun-20 | 65093 | 0.5269 | 532472 | 7067 | 62685 | 0.2163 | 268066 | 3530 |
| 6-Jun-20 | 66892 | 0.3857 | 537910 | 7149 | 65729 | 0.2004 | 271783 | 3581 |
| 7-Jun-20 | 69221 | 0.2621 | 543327 | 7229 | 68933 | 0.1798 | 275629 | 3632 |
| 8-Jun-20 | 72006 | 0.3858 | 548652 | 7307 | 72167 | 0.1452 | 279663 | 3683 |
| 9-Jun-20 | 75149 | 0.3379 | 553910 | 7384 | 75406 | 0.1593 | 283837 | 3734 |
| 10-Jun-20 | 78417 | 0.3518 | 559189 | 7460 | 78494 | 0.1943 | 288233 | 3786 |
| 11-Jun-20 | 81683 | 0.4282 | 564543 | 7536 | 81414 | 0.2358 | 292868 | 3840 |
| 12-Jun-20 | 84997 | 0.3672 | 569922 | 7611 | 84132 | 0.2626 | 297702 | 3897 |
| 13-Jun-20 | 88244 | 0.3817 | 575442 | 7685 | 86654 | 0.3523 | 302730 | 3956 |
| 14-Jun-20 | 91418 | 0.2618 | 581035 | 7759 | 88890 | 0.2784 | 308041 | 4018 |
| 15-Jun-20 | 94383 | 0.3852 | 586763 | 7834 | 90919 | 0.2422 | 313628 | 4083 |
| 16-Jun-20 | 97061 | 0.3720 | 592705 | 7910 | 92781 | 0.3714 | 319452 | 4151 |
| 17-Jun-20 | 99374 | 0.4487 | 598866 | 7987 | 94497 | 0.3459 | 325491 | 4222 |
| 18-Jun-20 | 101044 | 0.5157 | 605523 | 8066 | 95961 | 0.4193 | 331851 | 4297 |
| 19-Jun-20 | 101933 | 0.4554 | 612742 | 8146 | 97128 | 0.4933 | 338576 | 4376 |
| 20-Jun-20 | 102081 | 0.5496 | 620482 | 8229 | 98159 | 0.5175 | 345506 | 4458 |
| 21-Jun-20 | 101643 | 0.4445 | 628514 | 8316 | 98881 | 0.4490 | 352742 | 4543 |
| 22-Jun-20 | 100482 | 0.8345 | 636900 | 8410 | 99599 | 0.3751 | 360051 | 4632 |
| 23-Jun-20 | 98871 | 0.9648 | 645367 | 8511 | 100247 | 0.4159 | 367497 | 4726 |
| 24-Jun-20 | 96689 | 0.6972 | 653963 | 8619 | 100906 | 0.6824 | 375001 | 4824 |

## Notes

### Competing Interest Statement

The authors have declared no competing interest.

